# The impact of hormonal changes on Functional Neurological Disorder: An International Online Survey

**DOI:** 10.64898/2026.08.17.26360584

**Authors:** Laure von der Weid, Cristina Concetti, Ilaria Antonella Di Vico, Bettina Balint, Anita Barbey, Ilaria Bertaina, Jan Coebergh, Carlos Corral, Leandro da Costa, Lucas D’Andréa, Evdokia Efthymiou, Marialuisa Gandolfi, Axelle Gharib, Gabriela S. Gilmour, Daniela Kern, Richard A. Kanaan, Alex Lehn, Z Paige L’Erario, David D. G. Palmer, Petra Schwingenschuh, Catheline Stancu, Michele Tinazzi, Anne Weissbach, Ingrid Hoeritzauer, Selma Aybek

## Abstract

**Introduction:** Functional Neurological Disorder (FND) affects women approximately three times more often than men. This disparity has largely been attributed to higher trauma prevalence and diagnostic bias, while the potential contribution of hormonal influences has received little attention.

**Methods:** An online questionnaire was distributed through FND clinics in thirteen countries, assessing self-reported symptom change across five hormonal events: hormonal contraception, pregnancy, the menstrual cycle, menopause, and gender-affirming hormone therapy. Eligible participants were cisgender women with a diagnosis of FND, or gender minority individuals (transgender or non-binary). Perceived symptom change was rated on a five-point scale ranging from large improvement to large worsening.

**Results:** Among 262 respondents (96% female; mean age 39 years), several hormonal contexts were associated with self-reported symptom changes. Overall, hormonal contraception and pregnancy were frequently associated with worsening of motor and cognitive symptoms, and menopause with worsening across all symptom domains. Menstrual cycle analysis revealed a phase-dependent pattern: worsening was most frequently reported during menstruation and the luteal phase, whereas improvement was most frequent during the follicular phase. Reported changes were not uniform, with a substantial proportion of participants describing no change or improvement.

**Conclusion:** Self-reported FND symptom severity appears to vary with hormonal context, with motor and cognitive symptoms most consistently affected. Given the retrospective, self-report design, these findings are hypothesis-generating and support prospective research into the role of hormonal transitions in FND.

**What is already known on this topic:** FND affects women around three times more often than men, a disparity usually attributed to trauma exposure and diagnostic bias. Sex hormones modulate neural excitability and are established drivers of symptom variation in migraine, epilepsy and affective disorders, yet their relationship to FND symptom expression has not been systematically examined, leaving clinicians little evidence when patients ask how contraception, pregnancy or menopause might affect them.

**What this study adds:** We surveyed 2c2 people with FND internationally, who reported that symptoms varied with hormonal events, with motor and cognitive symptoms most consistently affected and a phase-dependent pattern across the menstrual cycle. Responses were heterogeneous rather than uniform, with a substantial proportion reporting no change or improvement in every context.

**How this study might affect research, practice or policy:** Hormonal context may warrant consideration in FND assessment, and transitions such as pregnancy and menopause may merit closer monitoring and recognition at onset of FND symptoms. The heterogeneity observed argues against prognostic generalisation to individual patients. Prospective studies with objective hormonal measures are needed to establish whether these associations reflect hormonal mechanisms, broader sensitivity to physiological change, or recall effects.

## Introduction

Functional Neurological Disorder (FND) is a common neurological disorder, affecting women two to three times more often than men[1]. This difference could partly be explained by the higher rate of psychological trauma and sexual assault in women [2,3]. However, women with a history of sexual abuse are still more at risk of developing FND than men with the same history [1,4], suggesting that additional biological, psychological, and/or societal factors contribute. Some of these factors could be sex hormones, as women are over-represented in all age groups except after 50 years old [5] and before puberty [6]. However, the literature has until recently offered no clear answer as to whether some or all sex hormones (e.g. estrogen, progesterone, or testosterone) influence FND symptomatology [7]. Emerging evidence now suggests that they do, and points specifically towards a potential influence of estrogen [8].

Beyond their reproductive roles, sex hormones act directly on the brain by crossing the blood-brain barrier and modulating neural excitability, neurotransmitter systems, and inflammatory pathways. Associations between neurological and psychiatric disorders and female sex hormones have been well documented [9–12]. Estrogen withdrawal is an established symptom trigger in other neurological disorders: menstrually related migraine affects 22% of women of childbearing age and is more severe than non-menstrual migraine [13,14], while 10–70% of menstruating women with epilepsy report catamenial seizures, which typically remit after menopause [15–17].

A comparable premenstrual pattern is seen in psychiatric conditions: 3–8% of women meet DSM-5 criteria for premenstrual dysphoric disorder [18], and less distinct premenstrual exacerbation has been described in anxiety, depression and schizophrenia [17,19–21]. Given the biopsychosocial nature of FND, hormonal fluctuations may similarly modulate symptom expression.

Gender-affirming hormone therapy constitutes a further hormonal event of interest, involving sustained exogenous estrogen, testosterone administration or blockade. Data on gender minority people with FND remain limited [22], though preliminary work suggests they are overrepresented in FND samples [9,23]. As these patients are predominantly assigned female at birth, hormone therapy alone is unlikely to explain this overrepresentation; minority stress has been proposed as a contributing factor [22].

Overall, there is a legitimate reason to explore whether sex hormones influence FND symptoms, yet existing evidence is limited. Hormonal transitions characterized by marked shifts in sex hormone levels, namely the menstrual cycle, pregnancy, menopause, hormonal contraception use, and gender-affirming hormone therapy, provide naturalistic frameworks within which to explore FND symptoms fluctuations associated with changes in hormone levels. The present study explores self-reported symptom changes during these transitions as a methodological lens to advance understanding of sex hormone influences on FND symptomatology. We hypothesize that (1) hormonal contraception (combined, or progesterone-based) would worsen symptoms, (2) pregnancy could also lead to a worsening of symptoms, (3) symptoms severity would fluctuate across the menstrual cycle, (4) menopause would improve symptoms and (5) gender-affirming hormone therapy would be associated with symptom changes according to the treatment received: improvement in case of testosterone and worsening in case of estrogens.

## Material and methods

Five naturalistic hormonal events were investigated: start of hormonal contraceptive use, pregnancy, the menstrual cycle, menopause, and hormonal replacement therapy in the context of a gender transition. Using custom-designed questionnaires administered through the REDCap platform, participants were asked to report symptom changes using a five-point Likert scale. Survey routing ensured that participants only completed questionnaires relevant to their personal hormonal history.

The participants signed a informed consent form. The protocol has been approved by the local ethics committee of the Canton of Vaud (BASEC number : 2024-00412) and followed the guidelines of the Declaration of Helsinki.

### Online survey

A specific questionnaire was associated with each hormonal event, assessing whether participants experienced changes in their FND symptoms during or following that event. Questionnaires were available in English, French, German, Italian, Spanish, and Portuguese. The complete questionnaires are available in **supplementary material**.

#### Symptom assessment

Participants evaluated symptom changes across five FND symptom categories: movement symptoms, seizures or convulsions, sensory symptoms and cognitive symptoms, as well as mood and anxiety, known to be implicated in FND [24] and sensitive to hormonal fluctuations. Responses were recorded on a five-point Likert scale: 1 = large improvement, 2 = slight improvement, 3 = no change, 4 = slight worsening, 5 = large worsening. Participants could also indicate that they did not experience a given symptom.

#### Demographic information and survey routing

At the beginning of the survey, participants provided demographic information including sex, gender identity, FND diagnosis details, and whether they had received a diagnosis of attention deficit hyperactivity disorder (ADHD) or autism spectrum disorder (ASD). An open field allowed them to report any additional comorbidities. Based on their responses, for instance, whether they had used hormonal contraception or experienced pregnancy, the relevant questionnaires were displayed automatically. Intersex identity and variations in sex characteristics were not specifically assessed.

#### Hormonal contraception

Participants who indicated current or past use of hormonal contraception were directed to the corresponding questionnaire, which assessed symptom changes during the period of contraceptive use. They were additionally asked to indicate the kind of contraception they used (e.g. Combined pill, progesterone-only pill) and the name of their contraceptive.

#### Pregnancy

Participants who were currently pregnant or had been pregnant in the past were directed to this section. They were asked to report any symptom changes experienced during pregnancy, irrespective of trimester, and were instructed to focus on their most recent pregnancy, if they had experienced more than one. For participants who had already given birth, an additional question assessed whether they had noticed symptom changes following the end of pregnancy.

#### Menstrual cycle

Participants were asked to self-report whether their menstrual cycle was regular or irregular. Participants then indicated whether they had noticed an improvement or a worsening during specific phases of their cycle: menstruation, the follicular phase, ovulation, and luteal phase.

#### Gender-affirming hormone therapy

Participants who had initiated or were currently undergoing gender-affirming hormone therapy were asked to report symptom changes since beginning that treatment. Hormonal fluctuations following gonadectomy were not specifically studied.

### Recruitment

Healthcare professionals working internationally with patients diagnosed with FND were contacted and asked to distribute a flyer to potential participants who met the inclusion criteria. Recruitment was primarily conducted through flyers that included a QR code linking directly to the online survey.

Further recruitment was supported through the French platform “Cap TNF” and the Swiss association “FNS Vereinigung Schweiz,”.

As quality control, we asked in the questionnaire for the name of the person who gave them the survey. This was to ensure that the survey had only been accessible to people who have a confirmed FND diagnosis.

The questionnaire was accessible between 07.08.2024 and 09.04.2026.

### Data cleaning and organization

All data were first visually inspected to identify and remove duplicate entries and potential aberrant values. Participants were then categorized by hormonal event (contraception, pregnancy, menstrual cycle, menopause, gender-affirming hormone therapy) and by symptom type (motor, functional/dissociative seizure, sensory, cognitive, mood, anxiety); a single participant could contribute to several events. For each analysis, participants whose FND symptoms began after the corresponding hormonal event were excluded, as no pre-event comparison was possible. Participants reporting an interval of fifteen years or more between the event and questionnaire completion were also excluded, since autobiographical recall of symptom change becomes substantially less reliable over longer intervals and this threshold retained most of the pregnancy and menopause reports while excluding the most remote recollections. Cisgender males were additionally excluded.

Concerning the menstrual cycle analysis, participants who indicated having a menstrual cycle that was “Very regular”, “Mostly regular” or “Sometime regular” were included in the sample. Participants with a cycle that was “Not regular” or “Chaotic” were excluded.

### Statistical analysis

Descriptive statistics were first calculated using both absolute and relative frequencies to summarize participants’ responses. For the questionnaires related to hormonal contraception, pregnancy, menopause, and gender transition, Wilcoxon signed-ranks were conducted to determine whether reported symptom changes differed from the midpoint (3 on the Likert scale), indicating no change. Additionally, correlations between all changes in symptoms and changes in mood and anxiety were assessed using Pearson correlations. Within each hormonal event, p-values across the symptom domains were adjusted for multiple comparisons using the false discovery rate procedure (FDR).

For the menstrual cycle questionnaire, a proportion test (*Z*-test) was used to assess whether one specific phase of the cycle was more frequently associated with changes in symptoms than others.

Descriptive statistics, graphics and inference statistics were performed in R studio (version 4.5.3).

## Results

### Demographics

A total of 262 participants signed the consent form. Most of the cohort were cisgender women (253, 96.56%) and 9 (3.5%) were part of a gender minority group. The mean age was 39 years. Overall, 17.24% had a diagnosis of ADHD, 10% of ASD and 5.24% with both ADHD and ASD. **Table 1**.

**Table 1.**
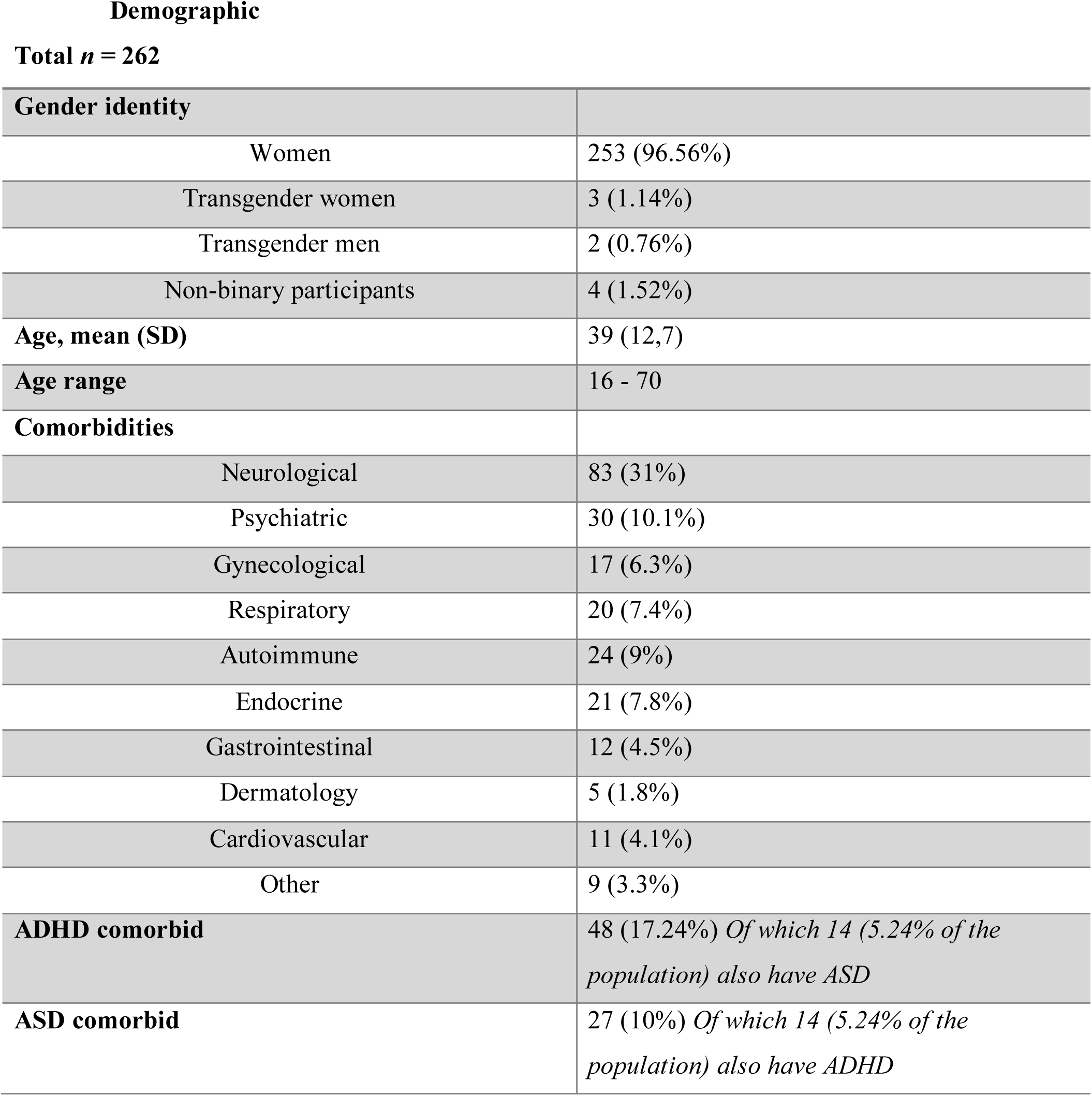
Gender distribution, age and comorbidities summary. Percentages reflect the proportion of the total sample. Age is presented as mean (standard deviation), with range from minimum to maximum age. A single participant may report multiple comorbidities and thus be counted in more than one category. All comorbidities were self-reported in an open question except ADHD and ASD that were explicitly asked.

**Table 2.**
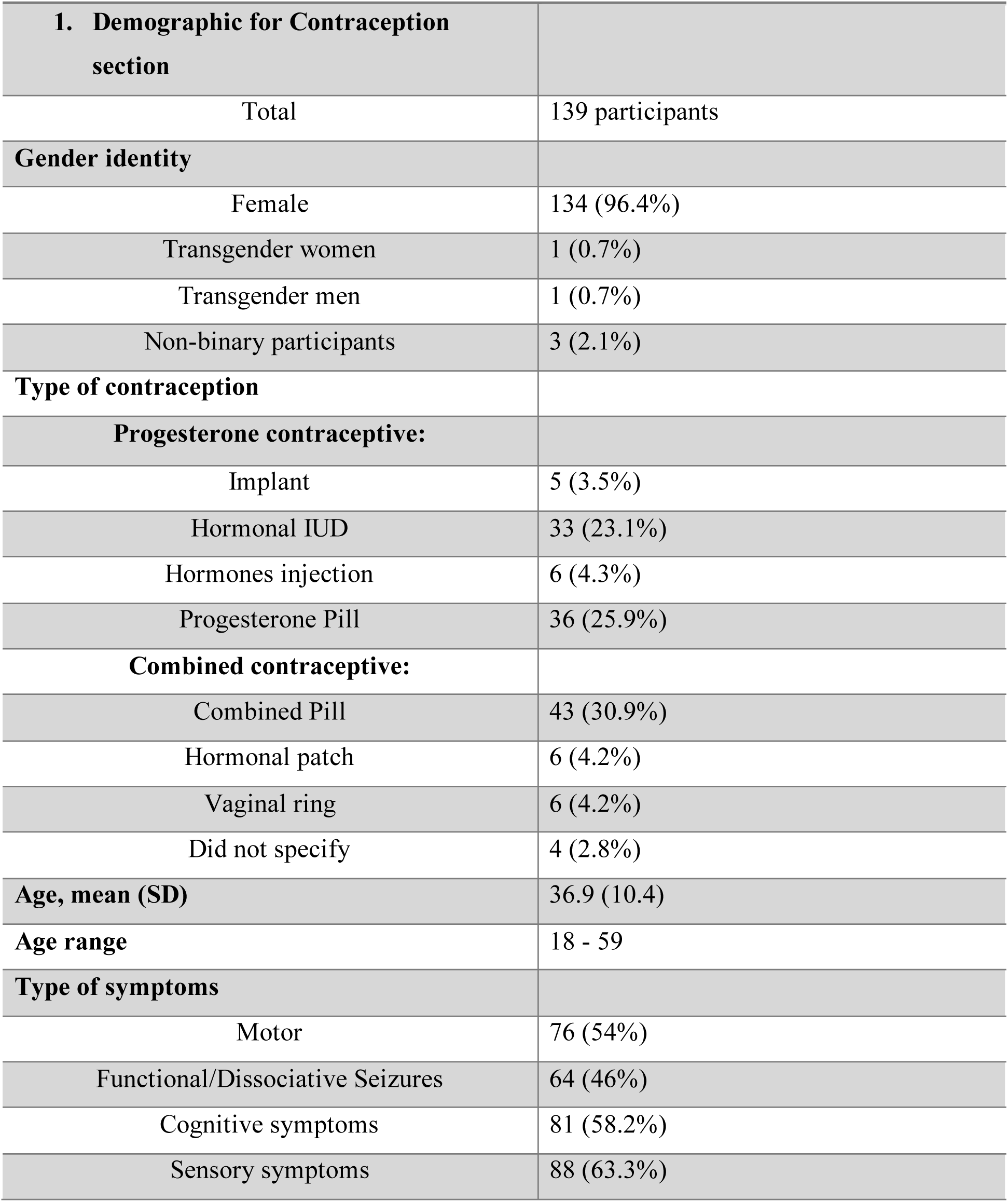
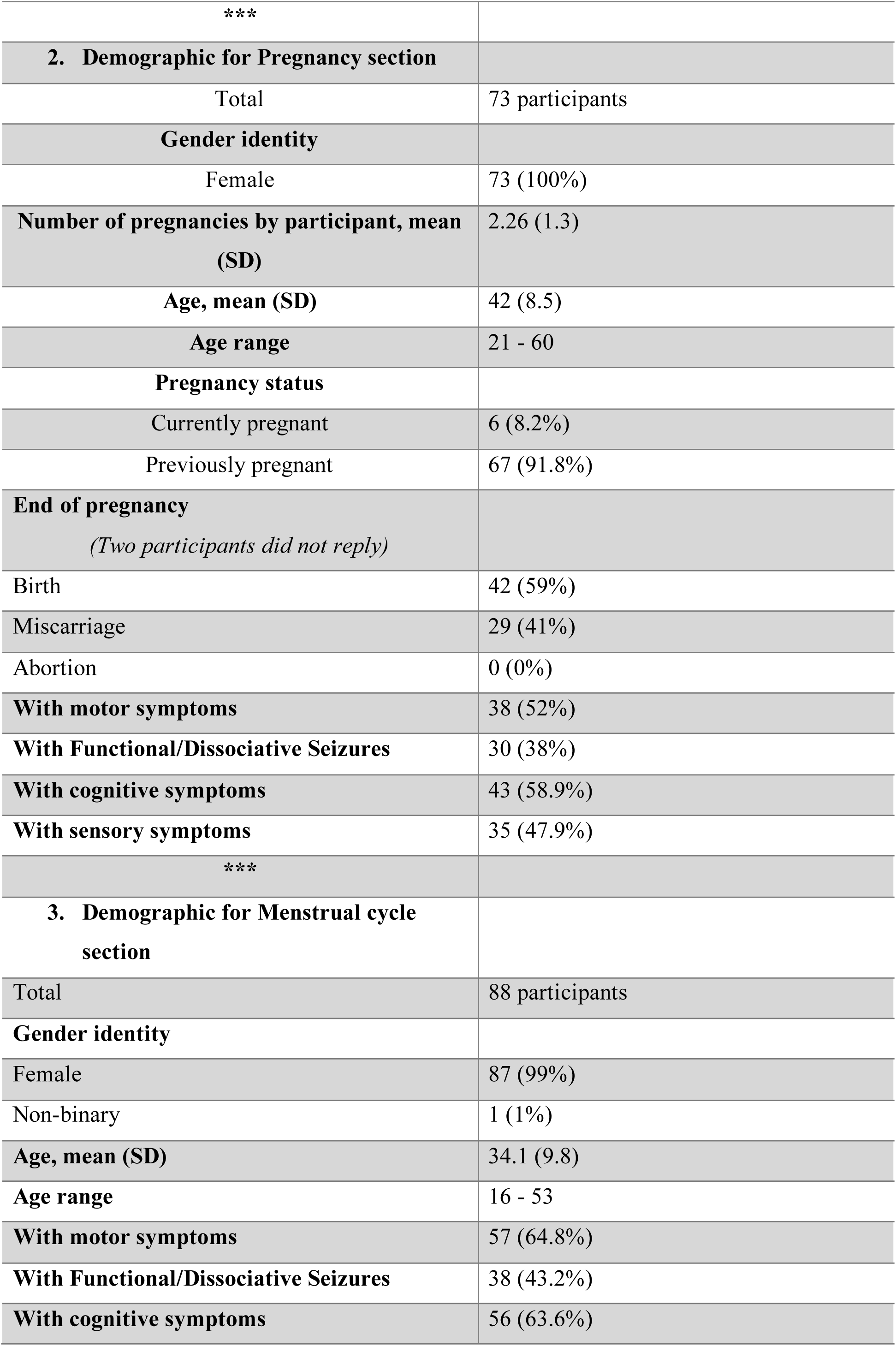
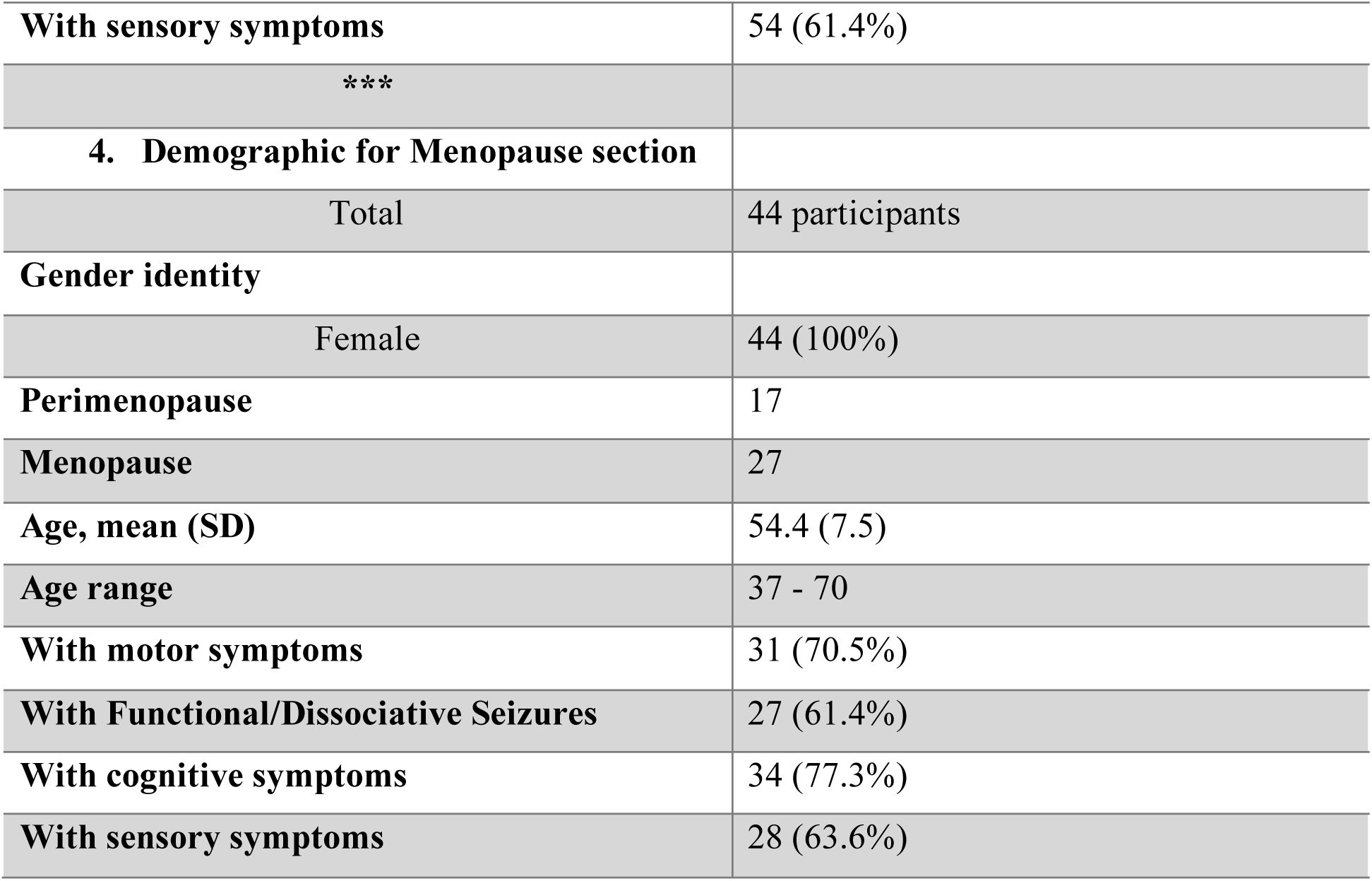
Participant characteristics by hormonal event. Demographic and clinical characteristics of participants contributing to each of the four hormonal events analysed (contraception, pregnancy, menstrual cycle, menopause). A single participant could contribute to more than one event, so subgroup totals are not mutually exclusive. Symptom rows show the number of participants within each subgroup reporting each FND symptom domain; participants could report multiple symptoms, so these do not sum to the subgroup total. Percentages are calculated within each event subgroup. Contraceptive types are shown as a percentage of hormonal contraceptive users. SD, standard deviation; IUD, intrauterine device.

The sample was geographically diverse, including participants from Europe, the Americas and Australia. **Fig 1**.

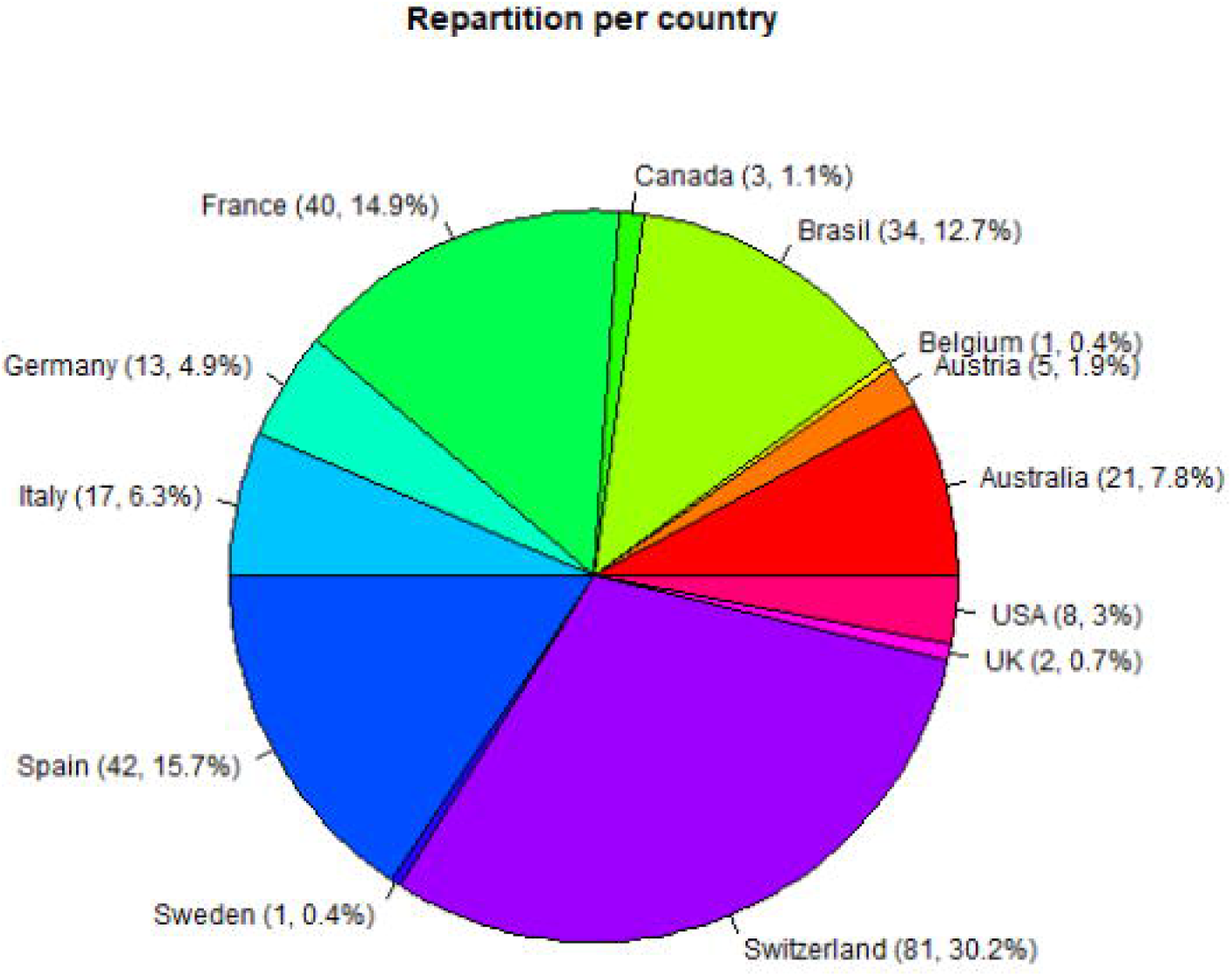

### Role of Contraception

Among participants reporting hormonal contraceptive use, motor symptoms worsened relative to the no-change midpoint (n = 76; Hodges–Lehmann median 3.5, 95% CI 3.0 to 4.5; p = 0.010, p_FDR = 0.040). Cognitive symptoms showed a similar direction of change that did not survive correction for multiple comparisons (n = 81; 3.5, 95% CI 3.0 to 4.5; p = 0.043, p_FDR = 0.086).

Functional/dissociative seizures (n = 64; 3.0, 95% CI 2.5 to 4.5; p = 0.61) and sensory symptoms (n = 68; 3.0, 95% CI 2.0 to 4.0; p = 1.0) showed no significant change. Across domains, reported change was consistently in the direction of worsening, reaching statistical significance only for motor symptoms after correction. **Fig 2**.

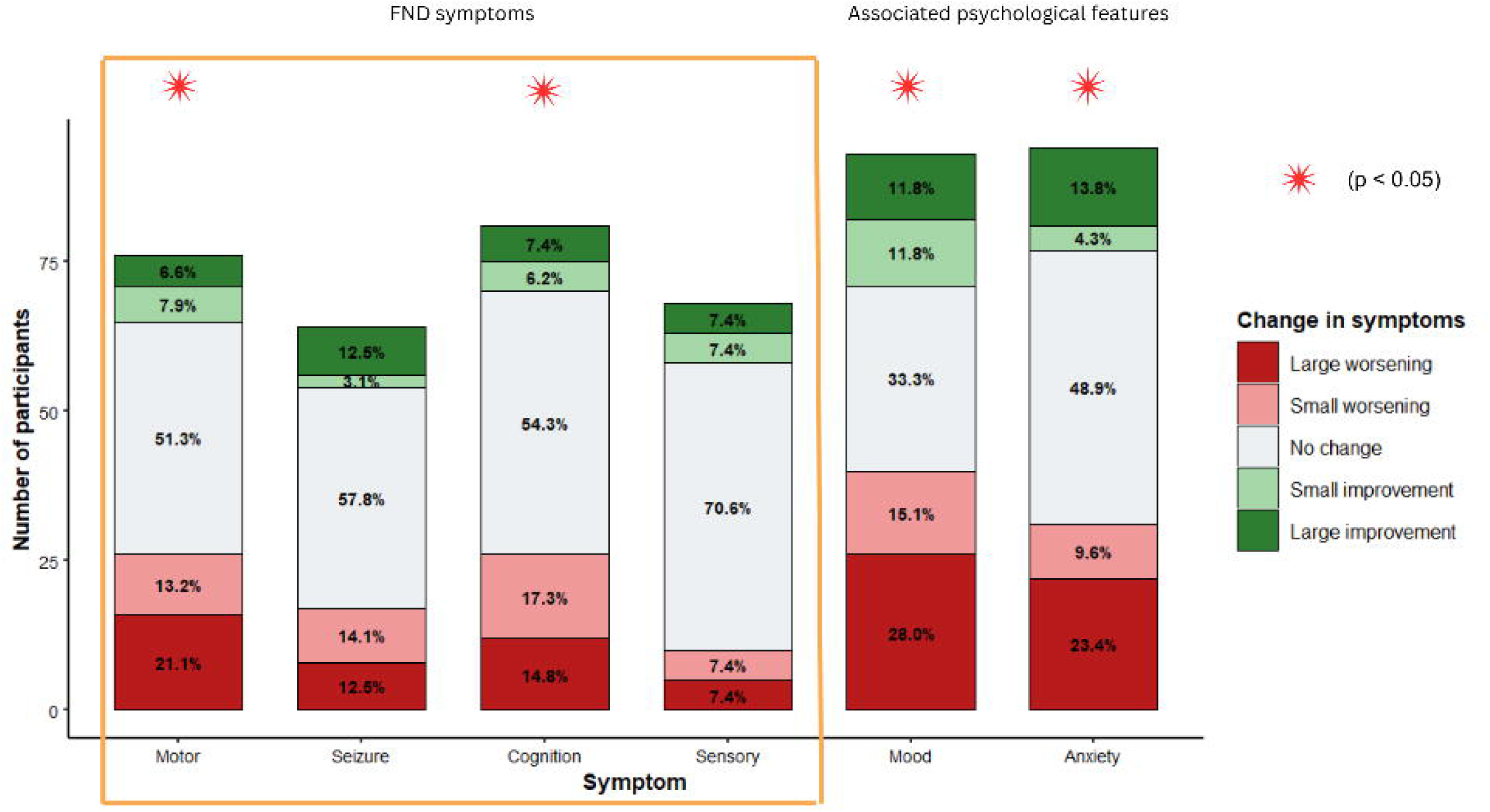

The result of progesterone and combined contraception separately are presented **in supplementary results (S1).**

### Pregnancy

#### Change in symptoms during pregnancy

Among participants who had been pregnant, motor and cognitive symptoms worsened relative to the no-change midpoint (motor: n = 37; Hodges–Lehmann median 4.5, 95% CI 3.0 to 4.5; p = 0.027, p_FDR = 0.040; cognitive: n = 42; 4.5, 95% CI 4.0 to 4.5; p = 0.003, p_FDR = 0.007). Functional/dissociative seizures (n = 29; 4.0, 95% CI 2.5 to 4.5; p = 0.28) and sensory symptoms (n = 34; 3.0, 95% CI 1.0 to 4.5; p = 0.72) showed no significant change. Mood and anxiety both worsened (mood: n = 44; 4.5, 95% CI 3.5 to 5.0; p_FDR = 0.007; anxiety: n = 40; 5.0, 95% CI 4.5 to 5.0; p_FDR = 0.001). **Fig 3 (A)**.

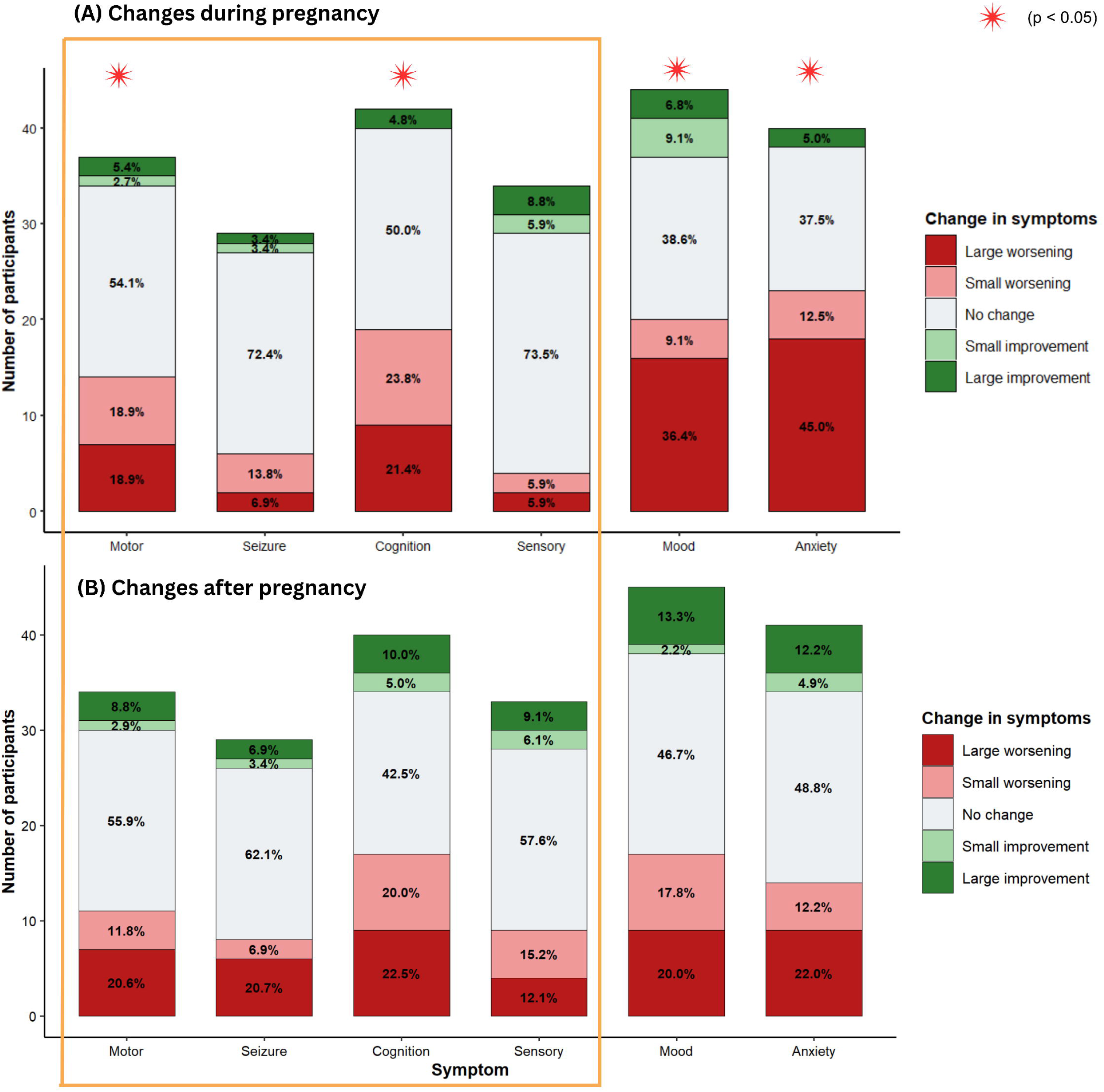

#### Change in symptoms after pregnancy

Following pregnancy, no symptom domain showed a significant change relative to the no-change midpoint after correction for multiple comparisons (motor: n = 34, Hodges–Lehmann median 4.0, 95% CI 3.0 to 5.0, p_FDR = 0.25; seizure: n = 29, 4.5, 95% CI 3.0 to 5.0, p_FDR = 0.25; cognitive: n = 40, 4.0, 95% CI 3.0 to 4.5, p_FDR = 0.25; sensory: n = 33, 3.5, 95% CI 2.5 to 4.5, p_FDR = 0.48). Mood and anxiety likewise showed no significant change (both p_FDR = 0.25). Although a substantial proportion of participants reported some change in each domain, no consistent direction emerged. **Fig 3 (B)**.

### Role of Menstrual cycle

Among participants with a regular menstrual cycle (N = 88, Table 4), approximately half reported symptom change across the cycle in every domain (motor 54.4%, functional/dissociative seizures 47.6%, cognitive 47.6%, sensory 48.1%). These changes were not uniformly distributed across cycle phases. Because subsample sizes within individual domains were small, worsening events were pooled across all four domains, yielding 152 observations from 88 participants. Pooled worsening deviated from an equal distribution (χ² = 25.6, df = 3, p < 0.001) and was most frequently reported during menstruation (n = 72, 47.4%) and the luteal phase (n = 45, 29.6%), and least during the follicular phase (n = 14, 9.2%). Pooled improvement showed the mirror pattern (χ² = 26.07, df = 3, p < 0.001), being most frequently reported during the follicular phase (n = 44) and at ovulation (n = 21). **Fig 4 (A)**.

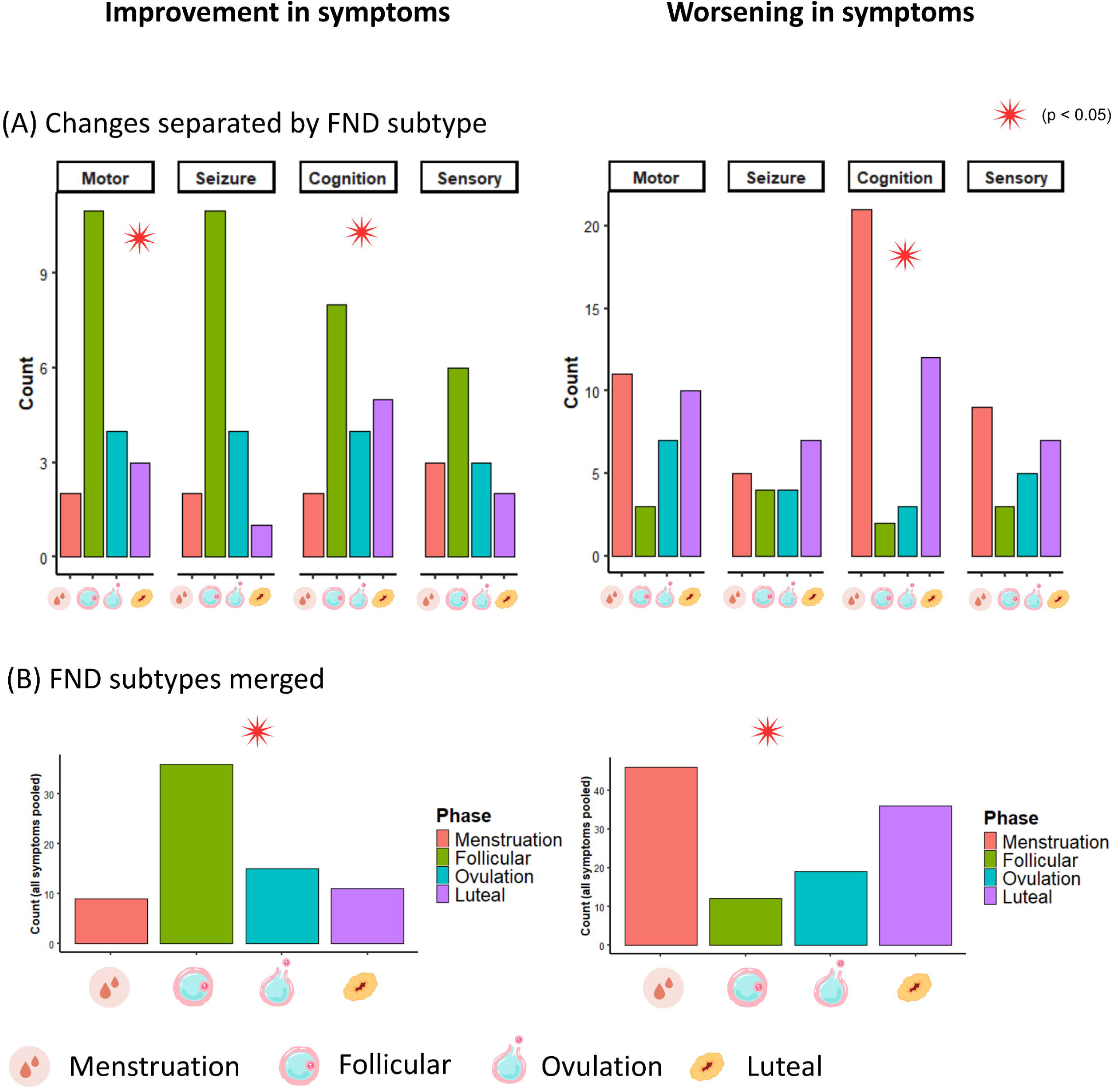

Analysed separately by domain, the same directional pattern was present throughout. After correction for multiple comparisons, worsening clustered in menstruation and the luteal phase for cognitive symptoms (χ² = 25.0, df = 3, p_FDR < 0.001, w = 0.81), while improvement clustered in the follicular phase for motor symptoms (χ² = 10.0, p_FDR = 0.035, w = 0.71) and functional/dissociative seizures (χ² = 13.6, p_FDR = 0.016, w = 0.87). Remaining domains showed the same direction without reaching significance. **Fig 4 (B)**. Full per-domain results are in Supplementary Table SX.

### Menopause

Among menopausal participants (n = 44, Table 5), symptoms worsened relative to the no-change midpoint across every domain: motor (n = 31; Hodges–Lehmann median 4.5, 95% CI 4.5 to 5.0; p_FDR < 0.001), functional/dissociative seizures (n = 27; 4.5, 95% CI 3.5 to 5.0; p_FDR = 0.008), cognitive (n = 34; 4.5, 95% CI 4.5 to 5.0; p_FDR < 0.001) and sensory symptoms (n = 28; 4.7, 95% CI 4.5 to 5.0; p_FDR = 0.001). Mood and anxiety also worsened markedly (mood: n = 33; 5.0, 95% CI 4.5 to 5.0; anxiety: n = 32; 5.0, 95% CI 4.5 to 5.0; both p_FDR < 0.001). More than half of participants reported small or large worsening in every domain. **Fig 5**.

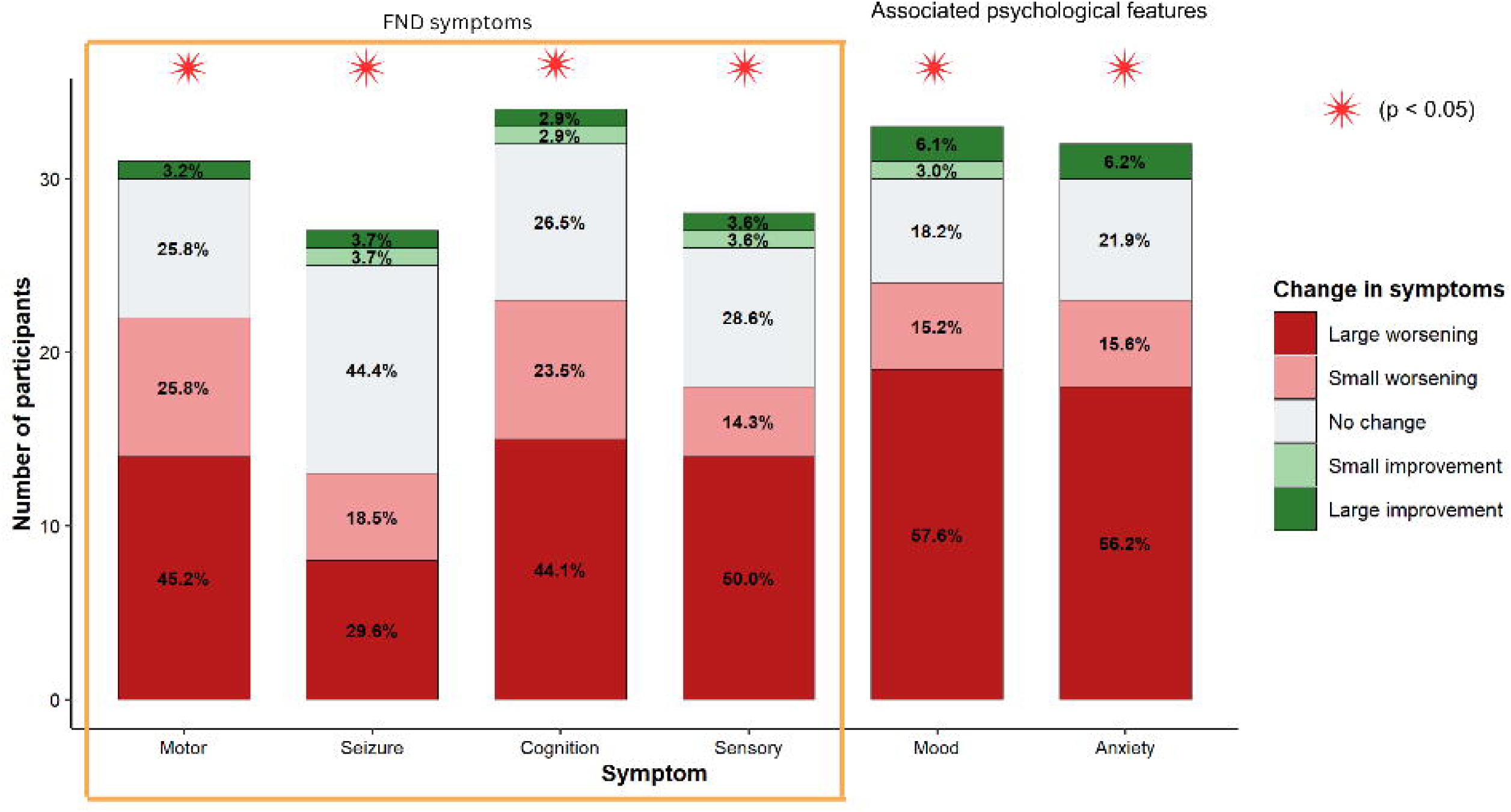

### Gender-affirming hormone therapy

In our sample, 6 participants reported receiving gender-affirming hormone therapy. Among this small number of participants, no symptom domain showed a statistically significant change after treatment onset. Overall, from this limited sample gender-affirming hormone therapy was not associated with significant worsening or improvement in motor, Functional/Dissociative seizure, cognitive or sensory symptoms. All participants reported improvement in mood and anxiety. Details are available in **supplementary materials (S5)**.

### Correlation between FND symptoms and psychological constructs

All FND symptoms changes have systematically been correlated with mood and anxiety changes. Across all symptoms and events, the correlation has been significant with a positive correlation **(the detail is available in supplementary material Table S2-S10)**.

## Conclusion

This study examined symptom change during hormonal contraceptive use, pregnancy, the menstrual cycle, menopause and gender-affirming hormone therapy. Overall, hormonal transitions showed a tendency toward symptom worsening across domains, reaching statistical significance for motor and cognitive symptoms. As the study was not powered by a priori and subsample sizes differed between domains, the absence of significance for functional/dissociative seizures and sensory symptoms should not be interpreted as an absence of effect, and some of the participants also showed improvements.

### Hormonal contraception leads to a relative increase of symptoms

Our findings suggest that hormonal contraception is associated with a worsening of symptoms in the motor domain. Additionally, cognitive symptoms also showed a statistically significant increase in severity, with worsening reported more frequently than improvement. In contrast, functional/dissociative seizures and sensory symptoms did not demonstrate a significant overall change, with reports of worsening and improvement more evenly distributed. Notably, even where overall effects were not statistically significant, approximately 30–50% of participants across symptom domains reported some degree of change, suggesting that hormonal contraception may influence symptom expression in a subset of individuals even when group-level effects are heterogeneous.

Hormonal contraception functions by suppressing LH and FSH release and inhibiting ovulation, resulting in consistently low estrogen levels, comparable to the early follicular phase, and progesterone levels near the limit of detection [25,26]. In psychiatric populations, findings have similarly been mixed: hormonal contraception has been linked to both increased and decreased depressive symptoms [27], with mood effects appearing to vary by formulation [28]. In our sample, mood and anxiety were also more likely to worsen following contraception initiation.

### FND symptoms fluctuate with the cycle

Symptom expression in FND varied across the menstrual cycle in a consistent direction. Across all symptom domains, worsening was most frequently reported during menstruation and the luteal phase, while improvement was most frequently reported during the follicular phase and ovulation. This pattern was present for both the combined analysis and, individually, for cognitive, motor and seizure symptoms.

The luteal phase and menstruation, periods of declining estrogen and progesterone, have been implicated in increased emotional reactivity [29], altered pain perception [30], and reduced stress resilience [31], all of which may exacerbate FND symptoms through their effects on central nervous system excitability, autonomic regulation, and affective processing. Conversely, the follicular phase, with its rising estrogen levels, appears to confer a degree of symptomatic relief, further supporting the hypothesis that estrogen may play a stabilizing role in FND.

#### Menopause increases symptoms severity

In our sample, menopause was associated with a marked worsening of symptoms across all domains, a pattern that distinguishes it from all other hormonal events examined in this study, where effects were more variable and domain-specific. This global impact across all FND symptoms, as well as mood and anxiety, suggests that menopause may represent a particularly vulnerable period for individuals with FND, potentially exerting a more pervasive impact on symptom expression than any other hormonal transition.

Unlike the cyclical shifts of the menstrual cycle, menopause entails sustained estrogen loss [32,33], which may act more pervasively on the systems implicated in FND.

#### Toward an estrogen hypothesis in FND

Across contraception, the menstrual cycle and menopause, low or declining estrogen was consistently associated with worsening and rising estrogen with improvement. This convergent pattern across independent hormonal contexts could suggest that estrogen may play a protective or stabilizing role in FND, while its withdrawal or sustained absence increases vulnerability to symptom expression. Finally, a consistent finding across all hormonal events was that changes in FND symptom severity were accompanied by corresponding changes in mood and anxiety. The nature of this association, however, cannot be determined from the present data. Three interpretations remain plausible: mood and anxiety changes may mediate the effect of hormonal fluctuations on FND symptoms; FND symptom worsening may itself precipitate affective deterioration; or hormones may exert parallel and independent effects on both symptom expression and affective states. Disentangling these pathways will require longitudinal designs with repeated hormonal and symptom measures. **Fig 6**.

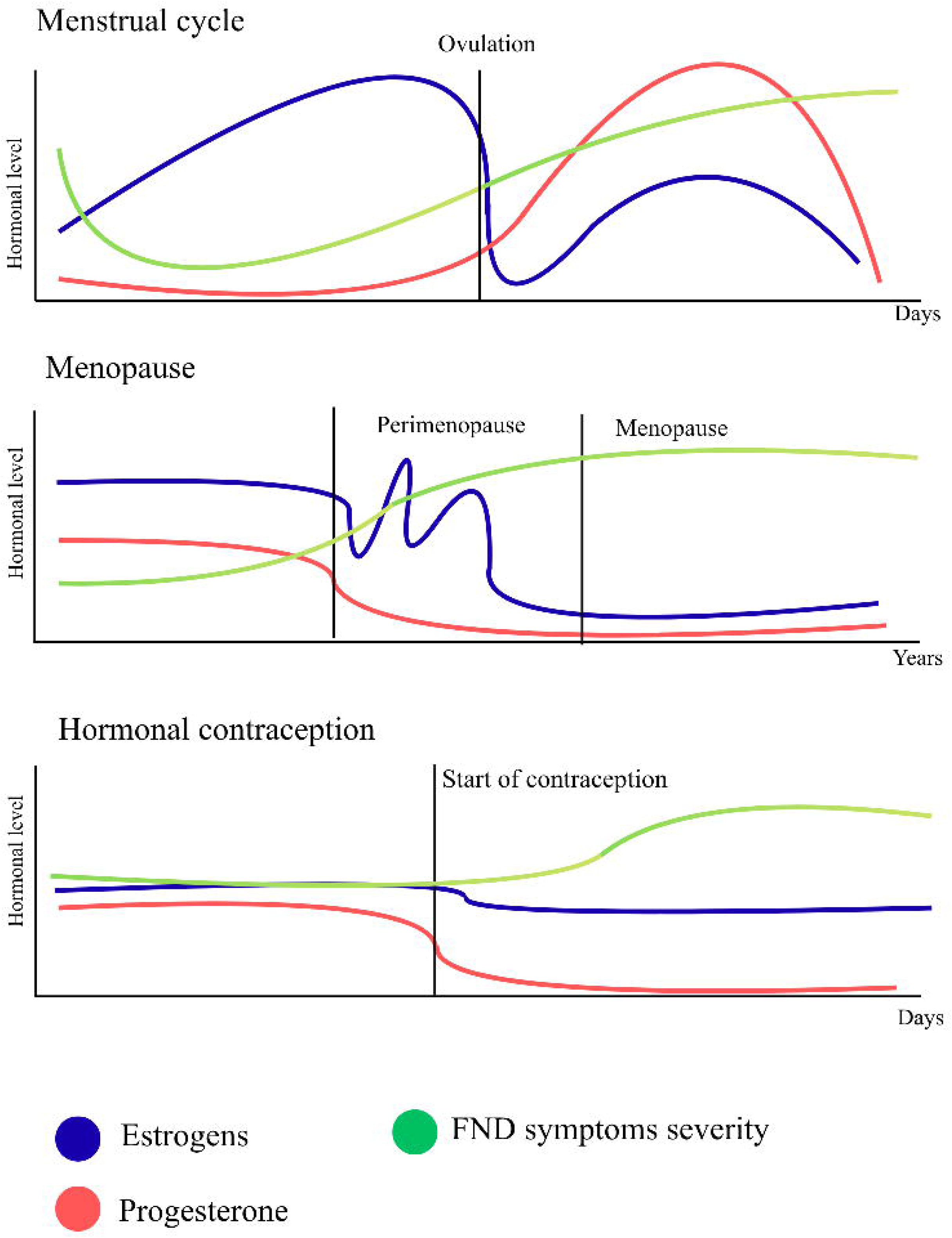

#### Pregnancy as a specific clinical context

In our sample, pregnancy was associated with significant worsening of motor and cognitive symptoms in 32.4% of participants, whereas functional/dissociative seizures did not change. This is an important clinical message, since it suggests pregnancy does not systematically increase seizure burden in FND. Half of participants reported no overall change. Despite pregnancy being a hyperestrogenic state, this worsening runs counter to the estrogen pattern seen elsewhere; in our sample mood and anxiety also worsened, and the physiological and psychological demands of pregnancy, rather than its hormonal profile, may drive symptom exacerbation. The postpartum period was more heterogeneous, though the proportion reporting major seizure worsening rose from 6% to 20%, suggesting a vulnerable subset.

#### Gender minorities are underrepresented in our sample

The limited number of participants receiving gender-affirming hormone therapy in our sample was low and this restricted the scope of the analyses that could be conducted. Recent studies have suggested a potential overrepresentation of gender minority individuals among patients with FND [22,23], a pattern that wasn’t observed in our sample.

Within our sample, participants receiving gender-affirming hormone therapy reported a tendency toward symptom improvement, although this reached significance only for mood-related outcomes. Future research with larger samples, or studies specifically designed to investigate gender minority populations, will be essential to better understand these relationships and to ensure that findings are both robust and inclusive.

### Limitations

This study has several limitations. First, participants were recruited through clinicians to reduce the risk of self-diagnosis. While this approach limits response bias, it cannot eliminate self-selection bias, as individuals who perceive a relationship between their hormonal status and FND may have been more inclined to participate. Although we cannot exclude the possibility that the survey link was shared externally, participants were required to provide the name of the referring clinician or institution, rendering fabricated access unlikely.

Second, the events examined were not corroborated by objective measures, such as serum hormone concentrations, and our outcomes reflect participants’ subjective, retrospective appraisals of change. Such reports are susceptible to recall bias and may not accurately capture objective symptom trajectories, particularly over extended intervals.

Third, we did not assess several factors known to vary across hormonal transitions and to influence FND symptom expression, most notably sleep disturbance and body weight. Both are affected by pregnancy and the menopausal transition and may act as confounders of the associations reported here.

### Key message

This study adds to the evidence that hormonal transitions are associated with variations in symptom expression in FND. Taken together, these findings support a biopsychosocial model in which hormonal transitions represent modulators of symptom variability in FND, rather than uniform causal factors. Clinically, these results underscore the importance of considering hormonal context in assessment and management, and suggest that periods such as pregnancy, the menstrual cycle, and menopause may warrant tailored support. Future research using longitudinal designs, objective hormonal measures and more diverse samples will be essential to clarify underlying mechanisms and to inform personalized approaches to care.

## Supporting information

Supplementary results

## Data Availability

All data produced in the present study are available upon reasonable request to the authors

## Acknowledgment

We thank all individuals that participated in the present study, among the professionals that actively distributed the survey as well as patients that filed the questionnaires.

## Bibliography

1 Kletenik I, Sillau SH, Isfahani SA, et al. Gender as a Risk Factor for Functional Movement Disorders: The Role of Sexual Abuse. Movement Disorders Clinical Practice. 2020;7:177–81. doi: 10.1002/mdc3.12863

2 Pan Y, Lin X, Liu J, et al. Prevalence of Childhood Sexual Abuse Among Women Using the Childhood Trauma Questionnaire: A Worldwide Meta-Analysis. *Trauma*, Violence & Abuse. 2021;22:1181–91.

3 Putica A, Agathos J, Felmingham K. How insights from posttraumatic stress disorder can inform treatment of functional neurological disorder. Nat Rev Psychol. 2025;4:654–68. doi: 10.1038/s44159-025-00479-1

4 Kletenik I, Holden SK, Sillau SH, et al. Gender disparity and abuse in functional movement disorders: a multi-center case-control study. J Neurol. 2022;269:3258–63. doi: 10.1007/s00415-021-10943-6

5 Baizabal-Carvallo JF, Jankovic J. Gender Differences in Functional Movement Disorders. Movement Disorders Clinical Practice. 2020;7:182–7. doi: 10.1002/mdc3.12864

6 Kanaan RA, Nicholson TR, Asan L, et al. Evidence for a diagnostic distinction between functional seizures and functional motor symptoms from the TriNetX electronic health record database. Psychological Medicine. 2026;56:e53. doi: 10.1017/S0033291726103456

7 Aybek S, Perez DL. Diagnosis and management of functional neurological disorder. BMJ. 2022;o64. doi: 10.1136/bmj.o64

8 Palmer DDG, Warren N, Morton A, et al. From Menarche to Menopause: Hormonal Influences on Functional Neurological Disorder. 2026;2026.07.16.26358260.

9 Bradlow R, Weid LVD, Zwickl S, et al. Why does FND mainly affect women? A consideration of gender imbalance in neuropsychiatric disease. Psychological Medicine. 2026;56:e104. doi: 10.1017/S0033291726104085

10 Psychiatric Symptoms Across the Menstrual Cycle… : Harvard Review of Psychiatry. Ovid. https://www.ovid.com/jnls/hrpjournal/fulltext/10.1097/hrp.0000000000000329∼psychiatric-symptoms-across-the-menstrual-cycle-in-adult (accessed 30 July 2026)

11 Roeder HJ, Leira EC. Effects of the Menstrual Cycle on Neurological Disorders. Curr Neurol Neurosci Rep. 2021;21:34. doi: 10.1007/s11910-021-01115-0

12 Czlonkowska A, Ciesielska A, Gromadzka G, et al. Estrogen and Cytokines Production - The Possible Cause of Gender Differences in Neurological Diseases. Current Pharmaceutical Design. 2005;11:1017–30. doi: 10.2174/1381612053381693

13 Vetvik KG, MacGregor EA, Lundqvist C, et al. Prevalence of menstrual migraine: A population-based study. Cephalalgia. 2014;34:280–8. doi: 10.1177/0333102413507637

14 Güven B, Güven H, Çomoğlu S. Clinical characteristics of menstrually related and non-menstrual migraine. Acta Neurol Belg. 2017;117:671–6. doi: 10.1007/s13760-017-0802-y

15 Reddy DS. The role of neurosteroids in the pathophysiology and treatment of catamenial epilepsy. Epilepsy Research. 2009;85:1–30. doi: 10.1016/j.eplepsyres.2009.02.017

16 Kumar D, Samar Iltaf S, Umer A, et al. The Frequency of Catamenial Epilepsy in Female Epileptic Patients of Reproductive Age Group Presented to the Tertiary Care Hospital. Cureus. 2020;12:e11635. doi: 10.7759/cureus.11635

17 Harden CL, Pulver MC, Ravdin L, et al. The Effect of Menopause and Perimenopause on the Course of Epilepsy. Epilepsia. 1999;40:1402–7. doi: 10.1111/j.1528-1157.1999.tb02012.x

18 Halbreich U, Borenstein J, Pearlstein T, et al. The prevalence, impairment, impact, and burden of premenstrual dysphoric disorder (PMS/PMDD). Psychoneuroendocrinology. 2003;28:1–23. doi: 10.1016/S0306-4530(03)00098-2

19 Lane T, Francis A. Premenstrual symptomatology, locus of control, anxiety and depression in women with normal menstrual cycles. Arch Womens Ment Health. 2003;6:127–38. doi: 10.1007/s00737-003-0165-7

20 Gonda X, Telek T, Juhász G, et al. Patterns of mood changes throughout the reproductive cycle in healthy women without premenstrual dysphoric disorders. Progress in Neuro-Psychopharmacology and Biological Psychiatry. 2008;32:1782–8. doi: 10.1016/j.pnpbp.2008.07.016

21 Ray P, Mandal N, Sinha VK. Change of symptoms of schizophrenia across phases of menstrual cycle. Arch Womens Ment Health. 2020;23:113–22. doi: 10.1007/s00737-019-0952-4

22 Lerario MP, Fusunyan M, Stave CD, et al. Functional neurological disorder and functional somatic syndromes among sexual and gender minority people: A scoping review. Journal of Psychosomatic Research. 2023;174:111491. doi: 10.1016/j.jpsychores.2023.111491

23 Bradlow RCJ, Zwickl S, Reardon P, et al. A systematic review of functional neurological disorder in transgender people. International Journal of Transgender Health. 2025;0:1–13. doi: 10.1080/26895269.2025.2538742

24 Pun P, Frater J, Broughton M, et al. Psychological Profiles and Clinical Clusters of Patients Diagnosed With Functional Neurological Disorder. Front Neurol. 2020;11. doi: 10.3389/fneur.2020.580267

25 Fleischman DS, Navarrete CD, Fessler DMT. Oral Contraceptives Suppress Ovarian Hormone Production. Psychol Sci. 2010;21:750–2. doi: 10.1177/0956797610368062

26 Jung-Hoffmann C, Heidt F, Kuhl H. Effect of two oral contraceptives containing 30 μg ethinylestradiol and 75 μg gestodene or 150 μg desogestrel upon various hormonal parameters. Contraception. 1988;38:593–603. doi: 10.1016/0010-7824(88)90044-3

27 Laird S, Ney LJ, Felmingham KL, et al. Hormonal Contraception and the Brain: Examining Cognition and Psychiatric Disorders. Current Psychiatry Research and Reviews. 2019;15:116–31. doi: 10.2174/1573400515666190521113841

28 Shahnazi M, Farshbaf Khalili A, Ranjbar Kochaksaraei F, et al. A Comparison of Second and Third Generations Combined Oral Contraceptive Pills’ Effect on Mood. Iran Red Crescent Med J. 2014;16:e13628. doi: 10.5812/ircmj.13628

29 Sundström Poromaa I, Gingnell M. Menstrual cycle influence on cognitive function and emotion processing—from a reproductive perspective. Front Neurosci. 2014;8. doi: 10.3389/fnins.2014.00380

30 Riley III JL, E. Robinson M, Wise EA, et al. A meta-analytic review of pain perception across the menstrual cycle. Pain. 1999;81:225–35. doi: 10.1016/S0304-3959(98)00258-9

31 Klusmann H, Luecking N, Engel S, et al. Menstrual cycle-related changes in HPA axis reactivity to acute psychosocial and physiological stressors – A systematic review and meta-analysis of longitudinal studies. Neuroscience & Biobehavioral Reviews. 2023;150:105212. doi: 10.1016/j.neubiorev.2023.105212

32 Santoro N. Perimenopause: From Research to Practice. Journal of Women’s Health. 2016;25:332–9. doi: 10.1089/jwh.2015.5556

33 Burger H, Woods NF, Dennerstein L, et al. Nomenclature and endocrinology of menopause and perimenopause. Expert Review of Neurotherapeutics. 2007;7:S35–43. doi: 10.1586/14737175.7.11s.S35

