## Supplementary material for "The impact of hormonal changes on Functional Neurological Disorder: An International Online Survey"

### Langage

Please complete the survey below.

Thank you!

---

To change langage, use tu upper right button

Pour changer la langue, utilisez le bouton en haut à droite

Um die Sprache zu ändern, verwenden Sie die Schaltfläche oben rechts

Per cambiare la lingua, utilizzare il pulsante in alto a destra

Para cambiar de idioma, utilice el botón situado en la parte superior derecha

Para mudar o idioma, utilize o botão situado na parte superior direita

### Consent form

Impact of sex hormones on the functional neurological disorder symptomatology
Principal Investigator: Selma Aybek

Sponsor: Selma Aybek

Good morning,

We would like to invite you to take part in our research project.

Your participation is entirely voluntary. All data collected as part of this project are subject to strict data protection regulations.

The research project is led by Selma Aybek, University of Freiburg. We will be happy to share the results with you.

To give you an overview of the project, here are the key points to bear in mind. More detailed information follows.

Why are we conducting this research project?
• Our research project aims to determine whether life events associated with hormonal change (start of hormonal contraception, pregnancy, menstrual cycle, menopause, gender transition) have an impact on the FND symptoms. We are looking for potential changes in quantity and quality of symptoms.

What do I have to do if I agree to take part?
• If you agree to take part in our project, you will be asked to answer different questionnaire about your symptoms, mood, anxiety. Different situations will be presented and if one or multiple situations apply to you, you will be able to indicate whether it affected your symptoms. You can leave the survey at any time without giving any explanation.
• The usual duration to complete the questionnaire doesn't exceed 50 minutes.

What are the benefits and risks of participating in this project?

Benefits
• Your participation in this research project will not bring you any direct benefit.
• By taking part, you are helping scientists to better understand how FND and sex hormones are linked. This would maybe lead to a better understanding of FND.
Risks and constraints
• There is no particular risk or constraints in this study.

By ticking the corresponding box at the end of the document, you certify that you have understood its contents and freely consent to take part in the project.

Detailed information

1. Aim of the project and selection of participants
In this information sheet, our research project is also simply referred to as the project. If you agree to take part, you are a project participant.

This project will enable us to examine the link between functional neurological disorder and life events associated with hormonal changes. We're asking you to get involved, because participation is open to anyone who received an FND diagnosis and is menstruated and/or use an hormonal contraception and/or has been pregnant and/or is menopausal and/or has started a gender transition.

2. General information on the project
A functional neurological disorder (FND) is a dysfunction of the nervous system in which no structural or organic lesion can be demonstrated. When someone suffers from FND, there are a multitude of signs by which the disease can manifest itself, such as muscular paralysis, uncontrolled tremors or cramps. In rarer cases, sensory impairments such as blindness, deafness or insensitivity may also occur. Diagnosing this complex disorder has always been a challenge for doctors. FND is generally treated with a combination of psychotherapy and physiotherapy.
Women are 2 to 3 times more at risk to develop FND than men. Today, there is no clear explanation for this gap. There is evidence that sex hormones have an influence on the nervous system and on many neurological diseases, such as epilepsy, and psychiatric disease, such as anxiety. However, we don't know in which way sex hormones influence the FND symptomatology and how life event (pregnancy, menopause) modulates the symptoms. Thus, there's a lack of knowledge on gender minority people (transgender, non-binary person) with FND. This study aims to collect information about the prevalence of gender minority people in FND population and about a potential link between gender transition and FND.
In this study, we'll collect information on how a list of life event have an impact on the symptoms, in the quantity and the quality. The listed events are: menstrual cycle, the intake of hormonal contraception, pregnancy, menopause or an hormonal treatment in the context of gender transition. Participants will be asked to indicate if they noticed any change in each symptoms before, during and after the event. Participants will only answer the questionnaires concerning events that they experienced. The total duration should not exceed 50 minutes.
This study is online and international. 318 participants will be recruited internationally.
We are carrying out this research project in accordance with the laws in force in Switzerland. We are also complying with all internationally recognised guidelines. The relevant ethics committee has examined and approved the research project.
The project is being carried out in compliance with Swiss legislation.

3. Project progress
If you agree to participate in this study, you will be directly redirected to the survey. You can take as much time you need, there is no time limit. Simply answer the question being as honest and detailed as you can. If you need to leave the survey, you will receive an unique code that will allow you to come back whenever you want. When the survey is completed, it will automatically be closed, and you don't have to do anything else.

4. Benefits
You will not derive any personal benefit from your participation.
If you take part in this research project, you will not receive any financial compensation. Participation in this research project does not entail any additional costs for you or your health insurance fund.
The results of this research project may contribute to a better understanding of the origin of SNF. It could also lead to new treatment options. You are not taking part in this research project.

5. Voluntary nature of participation and obligations
Your participation is entirely voluntary. If you choose not to take part, or if you choose to take part and change your mind during the course of the project, you will not have to justify your decision. This decision will have no adverse repercussions on your subsequent medical treatment.

6. Risks and constraints
There is no risks or constraints participating in this survey.

7. Results
No identifying information, such as your name or you address, will be collected. Consequently, there will not be any personnal results. All results will only be global results based on the total sample.

At the end of the project, the investigating doctor can send you a summary of the overall results.

8. Confidentiality of data and samples

8.1. Data processing and coding
As part of this research project, data relating to your person and your health will be collected and processed, in part automatically. This information is anonymized at the time of collection. Anonymization means that any data that might identify you (name, date of birth, etc.) is will not be collected. Consequently, it is not possible to link the data to your person..
Only a limited number of people may consult your data in unencrypted form, and then only in order to carry out the tasks required for the project. These people are bound by professional secrecy.

8.2. Protection of data and samples
All data protection guidelines are strictly adhered to. Your data may have to be transmitted in anonymized form, for example for publication, and may be made available to other researchers. The data collected will be downloaded in the form of a text file (.csv) for analysis and stored on the University of Fribourg's institutional server for a period of 10 years after the end of the collection.

8.3. Data protection in the event of re-use
Your data could later prove important in answering other questions and be sent to another database located in Switzerland or abroad to be used in other research projects. However, this database must comply with the same standards and requirements as the database for this project. By participating in this study, you agree that you anonymized could be reused in a future study.

8.4. Right of consultation during inspections
The project may be subject to inspections. These may be carried out by the relevant ethics commission or by the sponsor who initiated the project. In such cases, the physician-investigator must provide your data for the purposes of these inspections. All those involved are bound by the strictest professional secrecy.

9. Withdrawal of project
You may withdraw from the project at any time. However, medical data collected up to that point may still be analyzed in coded form.
In the event of withdrawal, your data and samples will continue to appear in coded form in project documents, primarily to ensure medical safety. You must therefore agree to this before giving your consent.

10. Remuneration
You will not receive any remuneration for your participation in this research project.

11. Liability
The University of Fribourg, which initiated the research project and is responsible for carrying it out, is liable for any damage you may suffer in connection with the project. The conditions and procedure are laid down by law. If you believe you have suffered a loss as a result of the study, please contact your investigator or your insurance company directly.

12. Financement

The project is fully funded by the University of Freiburg.

13. Contact person
You can ask questions about the project at any time. If you have any questions during or after the project, please contact :

Laure von der Weid


Prof. Dr. med. Selma Aybek
Université de Fribourg
Faculté des sciences et de médecine
Bureau 2.106, bâtiment PER09
Chemin du Musée 5
CH - 1700 Fribourg

Written declaration of consent for participation in a research project
Please read this form carefully. Do not hesitate to ask questions if you do not understand something or need clarification. Your written consent is required to participate in the project.

- I declare that I have been informed orally and in writing by the undersigned physician-investigator of the objectives and progress of the research project, as well as the possible advantages, disadvantages and risks.
- I am taking part in this project voluntarily and accept the contents of the information sheet I have been given on the above-mentioned project. I have had sufficient time to make my decision.
- I have received the answers to the questions I asked about taking part in this project. I will keep the information sheet and receive a copy of my written declaration of consent.
- I agree that the competent specialists from the project management and the relevant ethics committee may consult my uncoded data in order to carry out checks and inspections, provided that the confidentiality of this data is strictly guaranteed.
- I will be informed of any results that have a direct impact on my health. If I do not wish to receive this information, I will contact the investigating doctor.
- I am aware that my personal and health data (and samples) may be transmitted for research purposes as part of this project, and only in encrypted form. The sponsor ensures that data protection complies with Swiss standards and requirements.
- I may revoke my consent to take part in the project at any time and without having to give any reason, without this decision having any adverse repercussions on my further treatment. However, the data and samples collected up to the time of withdrawal will be analysed as part of the project.
- The data may be sent for analysis to another database in Switzerland or abroad, provided that it complies with standards and requirements that are at least equivalent to Swiss standards and requirements. All legal provisions relating to data protection are complied with.
- I am aware that the University of Fribourg is liable for any damage caused by the project.

I voluntarily agree to participate in this study. ◯ Yes ◯ No

Date and time ______________________

(Optional) If you are interested in the results of the study, you can provide an email address. We will send you the results when the study is done. Your address will be suppressed after. ______________________

### Demographic

This survey is a research study and is addressed to people who have a Functional Neurological Disorder diagnosis or have had symptoms in the past.

While science has made significant progress in understanding FND and its manifestations, little is known about the impact of hormones. We also don't exactly know how certain life stages, such as pregnancy or menopause, are affected by this disease.

The questionnaires are completely anonymous. No information about your identity will be collected. Please be as precise and detailed as you can.

The survey takes a maximum of 50 minutes. Thank you very much for your answers.

| | |
|---|---|
| Have you received a Functional Neurological Disorder diagnosis from a health professional? | ◯ Yes<br>◯ No |
| How did you receive this survey? | __________________________________ |
| When have you received this diagnosis? | ____________________________<br>(Please, enter the year you recieved this diagnosis. Ex: 2015) |
| What is the name of the person who gave you this survey? | ____________________________<br>(If you don't remember the name, please give the infos that you remember. Per ex, it was my neurologist in Brisbane's FND clinic) |
| Have you ever received a diagnosis of ADHD (Attention Deficit and/or Hyperactivity Disorder)? | ◯ Yes<br>◯ No |
| Have you ever received an ASD (Autism Spectrum Disorder) diagnosis? | ◯ Yes<br>◯ No |
| Do you have any other known chronic conditions? If yes, please specify which ones. | __________________________________ |
| What is your gender identity? | ◯ Female<br>◯ Male<br>◯ Female transgender (male to female)<br>◯ Male transgender (female to male)<br>◯ Non-binary<br>◯ Other |
| What is your age? | ____________________________<br>(Only number are allowed) |

Which country do you currently reside in?

__________________________________

### Contraception survey

This part of the survey focuses on hormonal contraception and its influence on FND symptoms.

If you are currently using or have used hormonal contraception in the past, try to remember how your symptoms evolved after starting the contraception, in comparison to the weeks before.

If you don't use any hormonal contraception and have never used it in the past, simply answer 'no' and skip to the next questionnaire.

| | |
|---|---|
| Do you use hormonal contraception?<br><br>For example: the pill, patch, implant, or hormonal IUD. | ◯ Yes<br>◯ No |
| Which type of contraception do you use? | ◯ Implant<br>◯ Hormonal IUD (intra-uterine dispositive)<br>◯ Hormones injection<br>◯ Estroprogestative pill<br>◯ Progestative pill<br>◯ Patch<br>◯ Vaginal Ring |
| If you know the brand or name of your contraception, please enter it in the response box. | __________________________________ |
| How many years have you been using this contraception? | __________________________________<br>(Enter the number of years. If your using this contraception for 3 years and 6 months, enter "3.5") |
| Have you used hormonal contraception in the past?<br><br>For example: the pill, patch, implant, or hormonal IUD. | ◯ Yes<br>◯ No |
| Which type of contraception did you use?<br><br>If you've used several types of contraception, please choose the last one you used. | ◯ Implant<br>◯ Hormonal IUD<br>◯ Hormones injection<br>◯ Estroprogestative pill<br>◯ Progestative pill<br>◯ Patch<br>◯ Vaginal Ring |
| How many years ago did you stop using this contraception?<br><br>Enter the number of years that have passed between when you stopped the contraception and now. | __________________________________<br>(Enter a number. Per exemple, if you stopped three years ago, enter "3") |

Did you observe any change in your symptoms when you started using your contraception?

If yes, please choose a rating between '1' and '5,' where '1' is a big improvement, '2' is a little improvement, '3' indicates no change, '4' suggests a little worsening, and '5' indicates a big worsening. If you need to specify, choose 'other.'

| | 1. I observed a large improvement in these symptoms. | 2. | 3. I didn't observe any change when I've started my contraception. | 4. | 5. I observed a large worsening in these symptoms. | I don't/did not experience these symptoms. | Other |
|---|---|---|---|---|---|---|---|
| With motor symptom or deficit (ex: dystonia, tremor, weakness) | ○ | ○ | ○ | ○ | ○ | ○ | ○ |
| With seizures or convulsions | ○ | ○ | ○ | ○ | ○ | ○ | ○ |
| With cognitive symptom (ex: memory, attention) | ○ | ○ | ○ | ○ | ○ | ○ | ○ |
| With sensory symptom or deficit | ○ | ○ | ○ | ○ | ○ | ○ | ○ |
| With vertigo and dizziness | ○ | ○ | ○ | ○ | ○ | ○ | ○ |
| With your everyday mood | ○ | ○ | ○ | ○ | ○ | ○ | ○ |
| With your everyday anxiety | ○ | ○ | ○ | ○ | ○ | ○ | ○ |

Motor symptom: Can you explain in you own words what "other" means to you? ______________________

Seizures or convulsions: Can you explain in you own words what "other" means to you? ______________________

Cognitive symptom: Can you explain in you own words what "other" means to you? ______________________

Sensory symptom: Can you explain in you own words what "other" means to you? ______________________

Vertigo symptom: Can you explain in you own words what "other" means to you? ______________________

Mood: Can you explain more what you're meaning with "other" ______________________

Anxiety: Can you explain more what you're meaning with "other" ______________________

Do you have any comments on the changes in your symptoms when you started hormonal contraception? ______________________

Do you have general comments on your symptomatology and your hormonal contraception? ______________________

Do you observe any change in your symptoms during the "pause" week (the week you are not taking your pill/patch/ring)? If yes, choose one button between "1" and "5" where "1" is a big improvement, "2" is a little improvement, "3" no change, "4" a little worsening and "5" a big worsening. If you need to specify, choose "other".

| | 1. I observed a large improvement in these symptoms. | 2. | 3. I didn't observe any change during the week without contraception. | 4. | 5. I observed a large worsening in these symptoms. | I don't/did not experience these symptoms. | other |
|---|---|---|---|---|---|---|---|
| With motor symptom or deficit (ex: dystonia, tremor, weakness) | ○ | ○ | ○ | ○ | ○ | ○ | ○ |
| With seizures or convulsions | ○ | ○ | ○ | ○ | ○ | ○ | ○ |
| With cognitive symptom (ex: memory, attention) | ○ | ○ | ○ | ○ | ○ | ○ | ○ |
| With sensory symptom or deficit | ○ | ○ | ○ | ○ | ○ | ○ | ○ |
| With vertigo and dizziness | ○ | ○ | ○ | ○ | ○ | ○ | ○ |
| With your everyday mood | ○ | ○ | ○ | ○ | ○ | ○ | ○ |
| With your everyday anxiety | ○ | ○ | ○ | ○ | ○ | ○ | ○ |

Motor symptoms or deficits: Can you explain with you own word what "other" means to you?

__________________________________

With seizures or convulsions: Can you explain with you own words what "other" means to you?

__________________________________

With cognitive symptoms: Can you explain in your own words what "other" means to you?

__________________________________

With sensory symptoms or deficit: Can you explain in your own words what "other" means to you?

__________________________________

With vertigo and dizziness: Can you explain in your own words what "other" means to you?

__________________________________

With mood: Can you explain more what you're meaning with "other"

__________________________________

With anxiety: Can you explain more what you're meaning with "other"

__________________________________

Do you have comments on any changes in your symptoms at different time in the month?

__________________________________

Do you have general comments concerning your mood, emotions, anxiety and your hormonal contraception?

__________________________________

### Pregnancy

This part of the survey focuses on pregnancy. If you have been pregnant multiple times, try to focus on your last pregnancy or the pregnancy you remember the most.

However, feel free to be as detailed as possible in the comments sections, especially if your symptoms were different in each pregnancy. If you've never been pregnant, simply answer "no" and skip to the next question.

| | |
|---|---|
| Are you currently pregnant? | ◯ Yes<br>◯ No |
| Please specify the number of months you are currently pregnant. | ______________________<br>(Enter a number. Per exemple: 3) |
| Have you ever been pregnant? | ◯ Yes<br>◯ No |
| Do you remember the year of your pregnancy?<br><br>(last pregnancy) | ______________________<br>(Ex: 2015) |
| Please specify the number of times you have been pregnant. | ______________________ |
| If you have been confronted with complications during the pregnancy, such as miscarriage or any concomitant condition, please provide details. If not, you can proceed to the next question. | ◯ Yes<br>◯ No<br>(In any of your pregnancies) |
| If you experienced complications during your pregnancy, please specify the kind of complication or complications. | ____________________________ |

**Did you observe any change in your symptoms during your pregnancy? If you've been pregnant multiple times, focus on your last pregnancy or the one you remember the most. Choose a rating between 1 and 5, where 1 is a big improvement, 2 is a little improvement, 3 indicates no change, 4 suggests a little worsening, and 5 indicates a big worsening. If you need to specify, choose 'other'.**

| | 1. I observed a large improvement in these symptoms. | 2. | 3. I didn't observe any change during my pregnancy. | 4. | 5. I observed a large worsening in these symptoms. | I don't/did not experience these symptoms | other |
|---|---|---|---|---|---|---|---|

| | | | | | | | |
|---|---|---|---|---|---|---|---|
| With motor symptom or deficit (ex: dystonia, tremor, weakness) | ○ | ○ | ○ | ○ | ○ | ○ | ○ |
| With seizures or convulsions | ○ | ○ | ○ | ○ | ○ | ○ | ○ |
| With cognitive symptom (ex: memory, attention) | ○ | ○ | ○ | ○ | ○ | ○ | ○ |
| With sensory symptom or deficit | ○ | ○ | ○ | ○ | ○ | ○ | ○ |
| With vertigo and dizziness | ○ | ○ | ○ | ○ | ○ | ○ | ○ |
| With your everyday mood | ○ | ○ | ○ | ○ | ○ | ○ | ○ |
| With your everyday anxiety | ○ | ○ | ○ | ○ | ○ | ○ | ○ |

---

Motor symptom: Can you explain in your own words what "other" means to you?

__________________________________

---

Seizures symptom: Can you explain in your own words what "other" means to you?

__________________________________

---

Cognitive symptom: Can you explain in your own words what "other" means to you?

__________________________________

---

Sensory symptom: Can you explain in your own words what "other" means to you?

__________________________________

---

Vertigo and dizziness: Can you explain in your own words what "other" means to you?

__________________________________

---

Mood: Can you explain with your own words what "other" means to you?

__________________________________

---

Anxiety: Can you explain with your own words what "other" means to you?

__________________________________

---

Do you have comments on any changes in your symptoms during my pregnancy?

__________________________________

---

Do you have general comments on your symptomatology and your pregnancy?

__________________________________

---

To what extent were your FND symptoms an obstacle during your pregnancy?

○ Not an obstacle at all
○ A little obstacle
○ A moderate obstacle
○ Very much
○ A severe obstacle

---

Do you have other general comments on your mood, anxiety, emotions and your pregnancy?

__________________________________

When did your last pregnancy end? (For example: 2018)

________________________________
(Enter a number)

How did your last pregnancy conclude?

- ◯ Birth
- ◯ Miscarriage
- ◯ Abortion
- ◯ Other

**Did you observe any change in your symptoms after your pregnancy ended? If you've been pregnant multiple times, focus on your last pregnancy or the one you remember the most. Choose a rating between 1 and 5, where 1 is a big improvement, 2 is a little improvement, 3 indicates no change, 4 suggests a little worsening, and 5 indicates a big worsening. If you need to specify, choose 'other'.**

| | 1. I observed a large improvement in these symptoms. | 2. | 3. I didn't observe any change after the end of my pregnancy. | 4. | 5. I observed a large worsening in these symptoms. | I don't/did not experience these symptoms. | other |
|---|---|---|---|---|---|---|---|
| With motor symptom or deficit (ex: dystonia, tremor, weakness) | ◯ | ◯ | ◯ | ◯ | ◯ | ◯ | ◯ |
| With seizures or convulsions | ◯ | ◯ | ◯ | ◯ | ◯ | ◯ | ◯ |
| With cognitive symptom (ex: memory, attention) | ◯ | ◯ | ◯ | ◯ | ◯ | ◯ | ◯ |
| With sensory symptom or deficit | ◯ | ◯ | ◯ | ◯ | ◯ | ◯ | ◯ |
| With vertigo and dizziness | ◯ | ◯ | ◯ | ◯ | ◯ | ◯ | ◯ |
| With your everyday mood | ◯ | ◯ | ◯ | ◯ | ◯ | ◯ | ◯ |
| with your everyday anxiety | ◯ | ◯ | ◯ | ◯ | ◯ | ◯ | ◯ |

Motor symptom: Can you explain with you own word what "other" means to you?

________________________________

Seizures or convulsion: Can you explain in your own words what "other" means to you?

________________________________

Cognitive symptom: Can you explain in your own words what "other" means to you?

________________________________

Sensory symptom: Can you explain with you own words what "other" means to you?

________________________________

Vertigo and dizziness: Can you explain with you own words what "other" means to you?

__________________________________

Mood: Can you explain more what "other" means to you?

__________________________________

Anxiety: Can you explain more what "other" means to you?

__________________________________

After the end of your pregnancy, have you:

(you can choose multiple answers, as long as it apply to your experience after the end of your pregnancy)

- ☐ Gained or lost a lot of weight quickly
- ☐ Slept more than necessary or had insomnia
- ☐ Been overwhelmed by events
- ☐ Been agitated or slowed down
- ☐ Felt useless or guilty
- ☐ Had difficulty concentrating
- ☐ Had suicidal thought

Do you have general comments concerning your symptoms, mood, emotions, anxiety at the end of your pregnancy?

__________________________________

Are-you breastfeeding? Or have you been breastfeeding by the past?

- ◯ Yes
- ◯ No

**Did you observe any change in your symptoms while breastfeeding? Choose one button between 1 and 5 where 1 is a big improvement, 2 is a little improvement, 3 no change, 4 a little worsening and 5 a big worsening. If you need to specify, choose "other".If you've been pregnant multiple time, focus on your last pregnancy or the pregnancy you remember the most.**

| | 1. I observed a large improvement in these symptoms. | 2. | 3. I didn't observe any change while breastfeeding. | 4. | 5. I observed a large worsening in these symptoms. | I don't/did not experience these symptoms. | Other: |
|---|---|---|---|---|---|---|---|
| With motor symptom or deficit (ex: dystonia, tremor) | ◯ | ◯ | ◯ | ◯ | ◯ | ◯ | ◯ |
| With seizures or convulsions | ◯ | ◯ | ◯ | ◯ | ◯ | ◯ | ◯ |
| With cognitive symptom (ex: memory, attention) | ◯ | ◯ | ◯ | ◯ | ◯ | ◯ | ◯ |
| With sensory symptom or deficit | ◯ | ◯ | ◯ | ◯ | ◯ | ◯ | ◯ |
| With vertigo and dizziness | ◯ | ◯ | ◯ | ◯ | ◯ | ◯ | ◯ |
| With your everyday mood | ◯ | ◯ | ◯ | ◯ | ◯ | ◯ | ◯ |
| with your everyday anxiety | ◯ | ◯ | ◯ | ◯ | ◯ | ◯ | ◯ |

Motor symptom: Can you explain with you own word what "other" means to you?

__________________________________

Seizures or convulsions: Can you explain with you own word what "other" means to you?

________________________________

Cognitive symptom: Can you explain with you own word what "other" means to you?

________________________________

Sensory symptom: Can you explain with you own word what "other" means to you?

________________________________

Vertigo and dizziness: Can you explain with you own word what "other" means to you?

________________________________

Mood: Can you explain in you own words what "other" means to you?

________________________________

Anxiety: Can you explain in your own words what "other" means to you?

________________________________

Do you have comments on functional neurological disorder and breastfeeding?

________________________________

Do you have more general comments on your experience of breastfeeding?

________________________________

### Menstrual cycle

This part of the survey focuses on your symptoms and your menstrual cycle.

If you have a menstrual cycle, meaning a monthly or close-to-monthly period, try to remember as well as you can the fluctuation in your symptoms during the month.

If you don't have a menstrual cycle, please answer the first question, and you will be redirected to the next questionnaire.

My menstrual cycle are :

- ◯ Very regular
- ◯ Mostly regular
- ◯ Sometime regular
- ◯ Not regular
- ◯ Chaotic
- ◯ I don't have menstrual cycle

Have you been diagnosed with POCS (polycystics ovary syndrome)?

- ◯ Yes
- ◯ No

Do you usually observe any worsening of your symptoms during your menstrual cycle? If need to specify, choose "other".

| | I observe a worsening of these symptoms during my periods. | I observe a worsening of these symptoms during the follicular phase (between day 1 of my period and around day 12). | I observe a worsening of these symptoms during ovulation (Between day 13 and 15). | I observe a worsening of these symptoms during the luteal phase (Between day 16 and 28). | I don't know/ I don't observe any change in these symptoms through my cycle. | I don't experience these symptoms. | Other |
|---|---|---|---|---|---|---|---|
| With motor symptom or deficit (ex: dystonia, tremor) | ◯ | ◯ | ◯ | ◯ | ◯ | ◯ | ◯ |
| With seizures or convulsions | ◯ | ◯ | ◯ | ◯ | ◯ | ◯ | ◯ |
| With cognitive symptom (ex: memory, attention) | ◯ | ◯ | ◯ | ◯ | ◯ | ◯ | ◯ |
| With sensory symptom or deficit | ◯ | ◯ | ◯ | ◯ | ◯ | ◯ | ◯ |
| With vertigo and dizziness | ◯ | ◯ | ◯ | ◯ | ◯ | ◯ | ◯ |

Motor symptom: Can you explain in you own words what "other" means to you?

__________________________________

Seizures or convulsions: Can you explain in you own words what "other" means to you?

__________________________________

Cognitive symtpom: Can you explain in you own words what "other" means to you?

__________________________________

Sensory symptom: Can you explain in you own words what "other" means to you?

__________________________________

Vertigo and dizziness: Can you explain in you own words what "other" means to you?

__________________________________

Do you usually observe any improvement of your symptoms during your menstrual cycle? If need to specify, choose "other".

| | I observe an improvement of these symptoms during my periods. | I observe an improvement of these symptoms during the follicular phase (between day 1 of my period and around day 12). | I observe an improvement of these symptoms during ovulation (Between day 13 and 15). | I observe an improvement of these symptoms during the luteal phase (Between day 16 and 28). | I don't know/I don't observe any change in these symptoms through my cycle. | I don't experience these symptoms. | Other. |
|---|---|---|---|---|---|---|---|
| With motor symptom or deficit (ex: dystonia, tremor, weakness) | ○ | ○ | ○ | ○ | ○ | ○ | ○ |
| With seizures or convulsions | ○ | ○ | ○ | ○ | ○ | ○ | ○ |
| With cognitive symptom (ex: memory, attention) | ○ | ○ | ○ | ○ | ○ | ○ | ○ |
| With sensory symptom or deficit | ○ | ○ | ○ | ○ | ○ | ○ | ○ |
| With vertigo and dizziness | ○ | ○ | ○ | ○ | ○ | ○ | ○ |

Motor symptom: Can you explain in you own words what "other" means to you?

__________________________________

Seizures or convulsions: Can you explain in you own words what "other" means to you?

__________________________________

Cognitive symptom: Can you explain in you own words what "other" means to you?

__________________________________

Sensory symptom: Can you explain in you own words what "other" means to you?

__________________________________

Vertigo and dizziness: Can you explain in you own words what "other" means to you?

__________________________________

Do you have comments on any changes in your symptoms during your menstrual cycle?

__________________________________

Do you have general comments on your symptomatology and your menstrual cycle?

__________________________________

### MEDI-Q

Please carefully review the list of provided symptoms. Please answer for each symptom that you have experienced during your periods in the last 12 months. If you did not experience a particular symptom, please answer "no" and skip to the next symptom on the list.

This questionnaire concerns the past year.

A. On average, in the past year on the days you had your period, did you... If you had this symptom, to what degree it interfered with your quality of life, your recreational or work activities, or your social relationships...

B. ...on days when you were menstruating? C. ...during the premenstrual phase (in the 7 days before the start of menstruation)? D. ...during the other days (outside the menstrual/premenstrual phase)?
Yes, more than half of the times I've had my period Yes, less than half the times I've had my period No (skip next question) 1. Not at all 2. A little 3. Moderately 4. Very much 1. Not at all 2. A little 3. Moderately 4. Very much 5. Never had this symptom during the premenstrual phase 1. Not at all 2. A little 3. Moderately 4. Very much 5. Never had this symptom during the other days

1. ...have pain in your lower abdomen? {yesmore1} {yesless1} {medi2} {medi26} {medi50}
2. ...have pain when urinating? {yesmore2} {yesless2} {medi3} {medi27} {medi51}
3. ...have pain during bowel movement? {yesmore3} {yesless3} {medi4} {medi28} {medi52}
4. ...have muscle/bone/joint pain? {yesmore4} {yesless4} {medi5} {medi29} {medi53}
5. ...feel bloated or did you experience breast tenderness? {yesmore5} {yesless5} {medi6} {medi30} {medi54}
6. ...experience nausea? {yesmore6} {yesless6} {medi7} {medi31} {medi55}
7. ...have headaches? {yesmore7} {yesless7} {medi8} {medi32} {medi56}
8. ...have digestive problems (heartburn, uncomfortable sense of fullness after meals ...)? {yesmore8} {yesless8} {medi9} {medi33} {medi57}
9. ...have diarrhea? {yesmore9} {yesless9} {medi10} {medi34} {medi58}
10. ...have constipation? {yesmore10} {yesless10} {medi11} {medi35} {medi59}
11. ...have discomfort due to vaginal bleeding (fear of stains or odors, discomfort from the tampon, difficulty or embarrassment during sexual activities...)? {yesmore11} {yesless11} {medi12} {medi36} {medi60}
12. ...have the feeling of being dirty? {yesmore12} {yesless12} {medi13} {medi37} {medi61}
13. ...feel excessively sad (easily crying, little drive to do things, loss of interest in usual activities ...)? {yesmore13} {yesless13} {medi14} {medi38} {medi62}
14. ...feel emotionally unstable (fluctuating mood, rapid transition from one mood to another even in response to minimal stimuli...)? {yesmore14} {yesless14} {medi15} {medi39} {medi63}
15. ...feel irritable or short-tempered (feeling nervous, not being able to bear unexpected events, people or situations, feeling angry easily...)? {yesmore15} {yesless15} {medi16} {medi40} {medi64}
16. ...feel impulsive (driven to act without thinking or planning...)? {yesmore16} {yesless16} {medi17} {medi41} {medi65}
17. ...feel anxious (agitated, tense, excessively insecure or indecisive, fearful that something bad could happen at any moment ...)? {yesmore17} {yesless17} {medi18} {medi42} {medi66}
18. ... excessively hungry (desire to overeat, loss of control over food...)? {yesmore18} {yesless18} {medi19} {medi43} {medi67}
19. ...feel a lack of hunger? {yesmore19} {yesless19} {medi20} {medi44} {medi68}
20. ...have insomnia (inability to fall or stay asleep)? {yesmore20} {yesless20} {medi21} {medi45} {medi69}
21. ...experience excessive sleepiness (sleeping during the day, not being able to get up in the morning ...)? {yesmore21} {yesless21} {medi22} {medi46} {medi70}
22. ...feel excessively tired (sluggish, with little energy...)? {yesmore22} {yesless22} {medi23} {medi47} {medi71}
23. ...have low sexual desire (reduced drive to have sexual activities, lack of sexual fantasies ...)? {yesmore23} {yesless23} {medi24} {medi48} {medi72}
24. ...have difficulty concentrating? {yesmore24} {yesless24} {medi25} {medi49} {medi73}

| | |
|---|---|
| Did you have sexual interactions that included vaginal penetration in the last year? | ◯ Yes<br>◯ No |

| | |
|---|---|
| On average, in the last year on the days you had your period, did you have pain during sexual interactions that included vaginal penetration? | ◯ Yes, more than half of the times I've had my period I had pain during vaginal penetration<br>◯ Yes, less than half of the times I've had my period I had pain during vaginal penetration<br>◯ No, I never had pain during vaginal penetration (end the questionnaire)<br>◯ I never had vaginal penetration on the days I had menstrual flow because I would have had too much pain<br>◯ I never had vaginal penetration on days when I had menstrual flow for reasons other than pain |
| On days when you were menstruating, to what degree this pain (or avoiding vaginal penetration) interfered with your quality of life, your recreational or work activities and your social relationships? | ◯ Not at all<br>◯ A little<br>◯ Moderately<br>◯ Very much |
| During the premenstrual phase (in the 7 days before the start of menstruation), if you had this symptom, to what degree it interfered with your quality of life, your recreational or work activities, or your social relationships? | ◯ Not at all<br>◯ A little<br>◯ Moderately<br>◯ Very much<br>◯ Never had this symptom during the premenstrual phase |
| During the other days (outside the menstrual/premenstrual phase), if you had this symptom, to what degree it interfered with your quality of life, your recreational or work activities and your social relationships? | ◯ Not at all<br>◯ A little<br>◯ Moderately<br>◯ Very much<br>◯ Never had this symptom during the other days |

### Menopause

This part of the survey focuses on menopause.

Try to remember as well as you can how your symptoms changed when you had the first signs of menopause. Feel free to be as detailed as possible in the comment section.

Are you in :

- ◯ Perimenopause (usually between 40 and 50 year old)
- ◯ Postmenopause (usually after 50 year old)
- ◯ I am not menopausal

How many years ago did you start your menopause?

__________________________
(Enter a number)

**Did you observe any change in your symptoms when you started menopause? Choose one button between 1 and 5 where 1 is a big improvement, 2 is a little improvement, 3 no change, 4 a little worsening and 5 a big worsening. If you need to specify, choose "other".**

| | 1. I observed a large improvement in these symptoms. | 2. | 3. I didn't observe any change when I've started menopause. | 4. | 5. I observed a large worsening in this symptom when I've started menopause. | I don't/did not experience these symptoms. | Other |
|---|---|---|---|---|---|---|---|
| With motor symptom or deficit (ex: dystonia, tremor, weakness) | ◯ | ◯ | ◯ | ◯ | ◯ | ◯ | ◯ |
| With seizures or convulsions | ◯ | ◯ | ◯ | ◯ | ◯ | ◯ | ◯ |
| With cognitive symptom (ex: memory, attention) | ◯ | ◯ | ◯ | ◯ | ◯ | ◯ | ◯ |
| With sensory symptom or deficit | ◯ | ◯ | ◯ | ◯ | ◯ | ◯ | ◯ |
| With vertigo and dizziness | ◯ | ◯ | ◯ | ◯ | ◯ | ◯ | ◯ |
| With your everyday mood | ◯ | ◯ | ◯ | ◯ | ◯ | ◯ | ◯ |
| With your everyday anxiety | ◯ | ◯ | ◯ | ◯ | ◯ | ◯ | ◯ |

Motor symptom: Can you explain in you own words what "other" means to you?

__________________________

Seizures or convulsions: Can you explain in you own words what "other" means to you?

__________________________

Cognitive symptom: Can you explain in you own words what "other" means to you?

__________________________

Sensory symptom: Can you explain in you own words what "other" means to you?

__________________________

| | |
|---|---|
| Vertigo and dizziness: Can you explain in you own words what "other" means to you? | ______________________ |
| Mood: Can you explain in your own words what "other" means to you? | ______________________ |
| Anxiety: Can you explain in your own words what "other" means to you? | ______________________ |
| Do you have comments on any changes in your symptoms when you started your menopause? | ______________________ |
| Do you have general comments on your symptomatology and menopause? | ______________________ |
| Do you have general comments concerning your mood, emotions, anxiety and menopause? | ______________________ |

### Transgender

The purpose of the next questions is to understand how feminizing/masculinizing hormones impact FND symptomatology. For these questions, try to remember as well as you can how your symptoms evolved after starting these hormones, focusing specifically on the impact of the hormones and not the entire gender transition.

| | |
|---|---|
| Are you currently undergoing hormone replacement therapy (HRT) as part of a gender transition, involving artificial hormones? | ◯ Yes<br>◯ No |
| Can you recall approximately at what age you first thought about a gender transition? If yes, please provide an approximate age. | ____________________<br>(Provide year and month if possible. Otherwise, explain in your own words.) |
| How many years ago did you officially start your gender transition? By 'officially,' we mean when you started using your current pronouns, name, or being socially accepted in your gender. | ____________________<br>(Enter a number) |
| How many years ago did you start hormone replacement therapy (HRT)? | ____________________ |
| Which hormones are you currently taking? (Estrogen, Testosterone, Other) | ____________________ |

**Did you observe any change in your symptoms when you started hormone replacement therapy, in comparison to when you started your social transition? Choose a rating between 1 and 5, where 1 is a big improvement, 2 is a little improvement, 3 indicates no change, 4 suggests a little worsening, and 5 indicates a big worsening. If you need to specify, choose 'other'.**

| | 1. I observed a large improvement in these symptoms. | 2. | 3. I didn't observe any change when I've started hormone replacement therapy. | 4. | 5. I observed a large worsening in these symptoms. | I don't/did not experience these symptoms. | Other |
|---|---|---|---|---|---|---|---|
| With motor symptom or deficit (ex: dystonia, tremor, weakness) | ◯ | ◯ | ◯ | ◯ | ◯ | ◯ | ◯ |
| With seizures or convulsions | ◯ | ◯ | ◯ | ◯ | ◯ | ◯ | ◯ |

| | | | | | | | |
|---|---|---|---|---|---|---|---|
| With cognitive symptom (ex: memory, attention) | ○ | ○ | ○ | ○ | ○ | ○ | ○ |
| With sensory symptom or deficit | ○ | ○ | ○ | ○ | ○ | ○ | ○ |
| With vertigo and dizziness | ○ | ○ | ○ | ○ | ○ | ○ | ○ |
| With your everyday mood | ○ | ○ | ○ | ○ | ○ | ○ | ○ |
| With your everyday anxiety | ○ | ○ | ○ | ○ | ○ | ○ | ○ |

---

Motor symptom: Can you explain in you own words what "other" means to you?

________________________________

---

Seizures or convulsions: Can you explain in you own words what "other" means to you?

________________________________

---

Cognitive symptom: Can you explain in you own words what "other" means to you?

________________________________

---

Sensory symptom: Can you explain in you own words what "other" means to you?

________________________________

---

Vertigo and dizziness: Can you explain in you own words what "other" means to you?

________________________________

---

Mood: Can you explain in you own words what "other" means to you?

________________________________

---

Anxiety: Can you explain in your own words what "other" means to you?

________________________________

---

Do you have comments on any changes in your symptoms when you started hormone replacement therapy?

________________________________

---

Do you have general comments on your symptomatology and your hormone replacement therapy?

________________________________

---

Do you have general comments concerning your mood, emotions, anxiety and your hormone replacement therapy?

________________________________

---

Do you have general comments on gender transition and functional neurological disorders?

________________________________
