## Supplementary results for "The impact of hormonal changes on Functional Neurological Disorder: An International Online Survey"

#### Role of Contraception

| Symptom | % Any Change | Large Worsening | Small Worsening | Large Improvement | Small Improvement | Mean Change | V | p-value (Wilcoxon) | p-adjusted (FDR) |
|---|---|---|---|---|---|---|---|---|---|
| Motor | 48.7% (37) | 21.1% (16) | 13.2% (10) | 6.6% (5) | 7.9% (6) | 3.34 | 517 | 0.01* | 0.034* |
| Seizure | 42.2% (27) | 12.5% (8) | 14.1% (9) | 12.5% (8) | 3.1% (2) | 3.11 | 210 | 0.61 | 0.733 |
| Cognitive | 45.7% (44) | 14.8% (12) | 17.3% (14) | 7.4% (6) | 6.3% (5) | 3.25 | 482 | 0.043* | 0.086 |
| Sensory | 29.4% (19) | 7.4% (5) | 7.4% (5) | 7.4% (5) | 7.4% (5) | 3.00 | 105 | 1.00 | 1.00 |
| Mood | 66.7% (62) | 28% (26) | 15.1% (14) | 11.8% (11) | 11.8% (11) | 3.35 | 1326 | 0.011* | 0.034* |
| Anxiety | 48.4% (30) | 22.6% (14) | 9.7% (6) | 9.7% (6) | 6.5% (4) | 3.29 | 745 | 0.09 | 0.136 |

*p < 0.05 = significant change

**Table S1. Change in FND symptoms following initiation of hormonal contraception (n = 139)** Among participants reporting hormonal contraceptive use, %reporting any change, and direction of change (large/small improvement/worsening). Mean change score (1=large improvement → 5=large worsening) and Wilcoxon test result.

#### Correlation between FND symptoms changes and associated psychological features

| FND symptoms | Mood | Anxiety |
|---|---|---|
| Motor | r = 0.5, P < 0.001 | r = 0.61, P < 0.001 |
| Seizure | r = 0.31, P = 0.013 | r = 0.47, P < 0.001 |
| Cognition | r = 0.6, P < 0.001 | r = 0.64, P < 0.001 |
| Sensory | r = 0.56, P < 0.001 | r = 0.61, P < 0.001 |

**Table S2. Correlation between change in FND symptoms with mood and anxiety after the start of an hormonal contraception**

### Dissociation between progesterone only and combined contraception

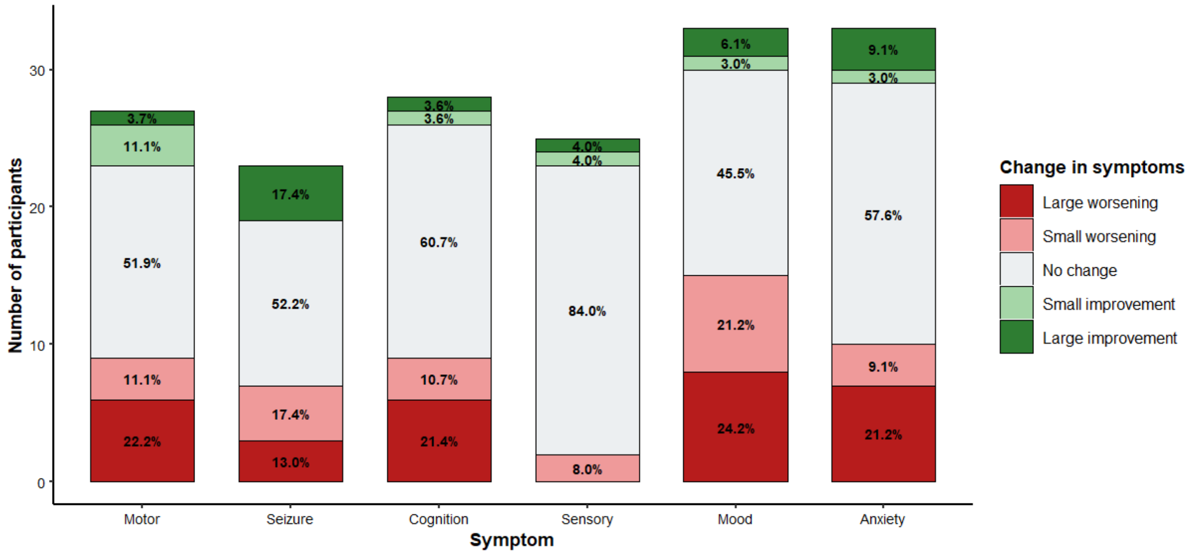


**Figure S1. Distribution of symptoms changes with progesterone only contraceptive**

In progesterone only contraceptive, motor symptoms showed no significant changes (V = 70.5, P = 0.0781, CI [2.99; 4.99]), Seizures showed no significant changes (V = 34, P = 0.9633, CI [2.49; 4.5], Cognition showed a significant worsening (V = 55.5, P = 0.043, CI [3.00; 5.00]), Sensory symptoms showed no significant changes (V = 4, P = 0.85, CI [1.99; 4.00]), Mood showed a significant change (V = 139, P = 0.016, CI[3.0; 4.5], Anxiety showed no significant change (V = 58.5, P = 0.121, CI[2.99; 5]).

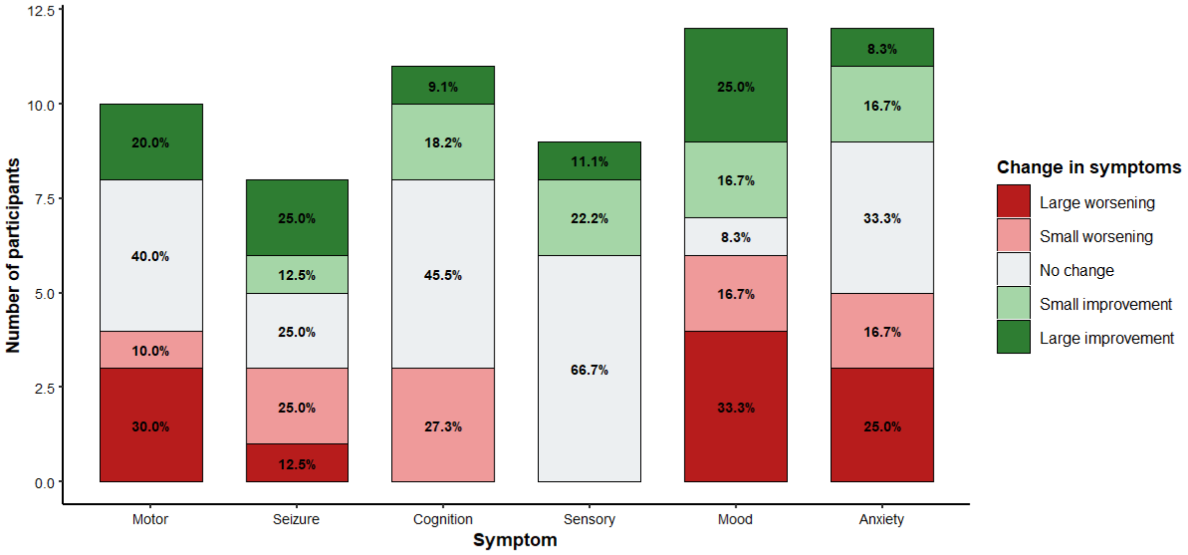


**Figure S2. Distribution of symptoms changes with combined contraceptive**

In combined contraceptive, motor symptoms showed no significant changes (V = 13, P = 0.656, CI [2.49; 5]), Seizures showed no significant changes (V = 9, P = 0.83, CI [1; 4.49], Cognition showed no significant worsening (V = 9, P = 0.824, CI [1.99; 4.00]), Sensory symptoms showed no significant changes (V = 0, P = 0.18, CI [2; 2]), Mood showed no significant change (V = 37, P = 0.74, CI[1.5; 4.5], Anxiety showed no significant change (V = 24.5, P = 0.388, CI[2.0 ; 4.99]).

#### Role of Pregnancy

#### During Pregnancy

| Symptom | % Any Change | Large Worsening | Small Worsening | Large Improvement | Small Improvement | Mean Change | V | p-value (Wilcoxon) | p-adjusted (FDR) |
|---|---|---|---|---|---|---|---|---|---|
| Motor | 44.7% (17) | 18.4% (7) | 18.4% (7) | 5.3% (2) | 2.6% (1) | 3.42 | 122 | 0.026* | 0.04* |
| Seizure | 26.7% (8) | 6.7% (2) | 13.1% (4) | 3.3% (1) | 3.3% (1) | 3.16 | 26 | 0.279 | 0.335 |
| Cognitive | 48.8% (22) | 20.9% (9) | 23.3% (10) | 4.7% (2) | 0% (0) | 3.58 | 199 | < 0.01* | < 0.01* |
| Sensory | 25.7% (26) | 8.6% (3) | 5.7% (2) | 5.7% (2) | 5.7% (2) | 2.94 | 19 | 0.715 | 0.715 |
| Mood | 60% (27) | 35.6% (16) | 8.9% (4) | 6.7% (3) | 8.9 (4) | 3.58 | 306 | < 0.01* | < 0.01* |
| Anxiety | 63.4% (26) | 46.3% (19) | 12.2% (5) | 4.9% (2) | 0% (0) | 3.95 | 294 | < 0.001* | 0.001 |

*p < 0.05 = significant change

**Table S3. Change in FND symptoms during pregnancy (n = 73)** Among participants reporting a pregnancy, % reporting any change, and direction of change (large/small improvement/worsening). Mean change score (1=large improvement → 5=large worsening) and Wilcoxon test result.

#### Correlation between FND symptoms changes and associated psychological features

| FND symptoms | Mood | Anxiety |
|---|---|---|
| Motor | r = 0.44, p = 0.007 | r = 0.49, P < 0.001 |
| Seizure | r = 0.23, P = 0.026 | r = 0.62, P < 0.001 |

| | | |
|---|---|---|
| Cognition | r = 0.46, P < 0.001 | r = 0.6, P < 0.001 |
| Sensory | r = 0.48, P < 0.001 | r = 0.51, P = 0.002 |

**Table S4. Correlation between change in FND symptoms with mood and anxiety during pregnancy**

### After Pregnancy

| Symptom | % Any Change | Large Worsening | Small Worsening | Large Improvement | Small Improvement | Mean Change | V | p-value (Wilcoxon) | p-adjusted (FDR) |
|---|---|---|---|---|---|---|---|---|---|
| Motor | 44.1% (15) | 20.6% (7) | 11.8% (4) | 8.8% (3) | 2.9% (1) | 3.32 | 85.5 | 0.14 | 0.254 |
| Seizure | 37.9% (11) | 20.7% (6) | 6.9% (2) | 6.9% (2) | 3.4% (1) | 3.31 | 49 | 0.15 | 0.254 |
| Cognitive | 57.5% (23) | 22.5% (9) | 20% (8) | 10% (4) | 5% (2) | 3.4 | 197 | 0.066 | 0.254 |
| Sensory | 42.4% (14) | 12.1% (4) | 15.2% (5) | 9.1% (3) | 6.1% (2) | 3.15 | 64 | 0.477 | 0.477 |
| Mood | 53.3% (24) | 20% (9) | 17.8% (8) | 13.3% (6) | 2.2% (1) | 3.29 | 193 | 0.2 | 0.254 |
| Anxiety | 51.2% (21) | 22% (9) | 12.2 (5) | 12.2 (5) | 4.9 (2) | 3.27 | 150 | 0.21 | 0.254 |

*p < 0.05 = significant change

**Table S5. Change in FND symptoms after pregnancy (n = 73)** Among participants reporting a pregnancy, % reporting any change, and direction of change (large/small improvement/worsening). Mean change score (1=large improvement → 5=large worsening) and Wilcoxon test result.

##### Correlation between FND symptoms changes and associated psychological features

| FND symptoms | Mood | Anxiety |
|---|---|---|
| Motor | r = 0.64, p < 0.001 | r = 0.70, p < 0.001 |
| Seizure | r = 0.70, p < 0.001 | r = 0.44, p = 0.018 |
| Cognition | r = 0.53, p < 0.001 | r = 0.40, p = 0.011 |
| Sensory | r = 0.61, p < 0.001 | r = 0.59, p < 0.001 |

**Table S6. Correlation between change in FND symptoms with mood and anxiety after pregnancy**

##### Most of participants did not see FND as an obstacle for a pregnancy

Sixty-six participants answered whether FND was an obstacle for their pregnancy or not. Among those, 46 (69%) did not see FND as an obstacle to their pregnancy. 7 (10%) thought it was a little obstacle, 5 (7.57%) a moderate obstacle and 8 (12.1%) a severe obstacle.

#### Role of Menstrual cycle

##### Worsening in symptoms during the menstrual cycle

Among participants with a regular menstrual cycle (N = 88, Table 4), symptom worsening was analysed separately by domain across the four cycle phases. For motor symptoms (n = 31), worsening did not deviate significantly from an equal distribution across phases ($\chi^2 = 5.00$, df = 3, p = 0.18, p_FDR = 0.36, w = 0.40), although it was descriptively more frequent during menstruation (n = 11) and the luteal phase (n = 10) than during the follicular phase (n = 3). For functional/dissociative seizures (n = 20), no significant phase-related pattern was observed ($\chi^2 = 1.20$, df = 3, p = 0.84, p_FDR = 0.84, w = 0.24), with worsening distributed across phases without a clear peak. Cognitive symptoms (n = 38) showed a significant phase effect that survived correction ($\chi^2 = 24.95$, df = 3, p_FDR < 0.001, w = 0.81), with worsening reported predominantly during menstruation (n = 21) and the luteal phase (n = 12) and rarely during the follicular phase (n = 2). Sensory symptoms (n = 24) did not differ significantly across phases ($\chi^2 = 4.46$, df = 3, p = 0.37, p_FDR = 0.49, w = 0.37), though worsening again occurred descriptively more often during menstruation (n = 9) and the luteal phase (n = 7) than the follicular phase (n = 3).

| **Symptom** | **% Any Change** | **Worsening during periods** | **Worsening during follicular** | **Worsening during ovulation** | **Worsening during luteal** | **p-value (Chi-squared)** | **p-adjusted (FDR)** |
|---|---|---|---|---|---|---|---|
| Motor | 54.4% (31) | 19.3% (11) | 5.3% (3) | 12.3% (7) | 17.5% (10) | 0.172 | 0.36 |
| Seizure | 47.6% (16) | 11.9% (5) | 9.5% (4) | 9.5% (4) | 16.7% (7) | 0.753 | 0.24 |
| Cognitive | 67.9% (38) | 37.5% (21) | 3.6% (2) | 5.4% (3) | 21.4% (12) | < 0.001* | < 0.001* |
| Sensory | 48.1% (26) | 18.5% (10) | 15.6% (3) | 9.3% (5) | 14.8% (8) | 0.343 | 0.489 |

*p < 0.05 = significant change

**Table S7. Change in FND symptoms during the menstrual cycle (n = 88)** Among participants reporting a menstrual cycle, % reporting each symptom, % reporting any change.

#### Improvement in symptoms during the menstrual cycle

Improvement was analysed separately for each symptom domain across cycle phases. Motor symptoms improved in 36.4% of participants, most often during the follicular phase (20%, n =

11), a distribution that deviated significantly from equal after correction ($\chi^2 = 10.0$, df = 3, p_FDR = 0.035, w = 0.71). Functional/dissociative seizures improved in 40.9% of participants, again predominantly during the follicular phase (25%, n = 11), and also reached significance ($\chi^2 = 13.56$, df = 3, p_FDR = 0.016, w = 0.87). Cognitive symptoms improved in 38.8% of participants but showed no significant phase distribution ($\chi^2 = 3.95$, df = 3, p_FDR = 0.41), with improvements reported during the follicular (16.3%) and luteal (10.2%) phases. Sensory symptoms improved in 26.8% of participants, with no significant phase effect ($\chi^2 = 2.33$, df = 3, p_FDR = 0.58).

When all symptoms were combined a chi-squared goodness-of-fit test indicated a significant effect of cycle phase ($\chi^2 = 28.43$, df = 3, $p < 0.001$). Improvements were most frequently reported during the follicular phase (n = 44), and the ovulation phase (n = 21).

| **Symptom** | **% Any Change** | **Improvement during periods** | **Improvement during follicular** | **Improvement during ovulation** | **Improvement during luteal** | **p-value (Chi-squared)** | **p-adjusted (FDR)** |
|---|---|---|---|---|---|---|---|
| Motor | 36.4% (20) | 3.6% (2) | 20% (11) | 7.3% (4) | 3% (5.5) | 0.018* | 0.036* |
| Seizure | 40.9% (18) | 4.5% (2) | 25% (11) | 9.1% (4) | 2.3% (1) | 0.003* | 0.016* |
| Cognitive | 38.8% (19) | 4.1% (2) | 16.3% (8) | 8.2% (4) | 10.2% (5) | 0.267 | 0.407 |
| Sensory | 26.8% (15) | 5.4% (3) | 10.7% (6) | 7.1% (4) | 3.6% (2) | 0.506 | 0.577 |

*p < 0.05 = significant change

**Table S8. Change in FND symptoms during the menstrual cycle (n = 88)** Among participants reporting hormonal contraceptive use, % reporting each symptom, % reporting any change

#### Role of Menopause

| Symptom | % Any Change | Large Worsening | Small Worsening | Large Improvement | Small Improvement | Mean Change | V | p-value (Wilcoxon) | p-adjusted (FDR) |
|---|---|---|---|---|---|---|---|---|---|
| Motor | 63.6% (23) | 45.2% (14) | 25.8% (8) | 3.2% (1) | 0% (0) | 4.1 | 260 | < 0.001* | < 0.001* |
| Seizure | 55.5% (15) | 29.6% (8) | 18.5% (5) | 3.7% (1) | 3.7% (1) | 3.66 | 106 | < 0.01* | < 0.01* |
| Cognitive | 73.5% (25) | 44.1% (15) | 23.5% (8) | 2.9% (1) | 2.9% (1) | 4.02 | 302 | < 0.001* | < 0.001* |
| Sensory | 71.4% (20) | 850% (14) | 14.3% (4) | 3.6% (1) | 3.6% (1) | 4.03 | 194 | < 0.001* | 0.001* |
| Mood | 11.6% (6) | 57.6% (19) | 15.2 % (5) | 6.1% (2) | 3% (1) | 4.15 | 340 | < 0.001* | < 0.001* |
| Anxiety | 78.1% (7) | 56.2% (18) | 15.6% (5) | 6.2% (2) | 0% (0) | 4.16 | 294 | < 0.001* | < 0.001* |

*p < 0.05 = significant change

**Table S9. Change in FND symptoms during or after menopause (n = 44)** Among participants reporting being menopausal, % reporting any change, and direction of change (large/small improvement/worsening). Mean change score (1=large improvement → 5=large worsening) and Wilcoxon test result.

#### Correlation between FND symptoms changes and associated psychological features

| FND symptoms | Mood | Anxiety |
|---|---|---|
| Motor | r = 0.54, P < 0.01 | r = 0.66, P < 0.001 |
| Seizure | r = 0.48, P = 0.011 | r = 0.49, P < 0.01 |
| Cognition | r = 0.48, P < 0.01 | r = 0.48, P < 0.01 |
| Sensory | r = 0.61, P < 0.001 | r = 0.57, P < 0.01 |

**Table S10. Correlation between change in FND symptoms with mood and anxiety during menopause**

#### Distinction between menopause and perimenopause

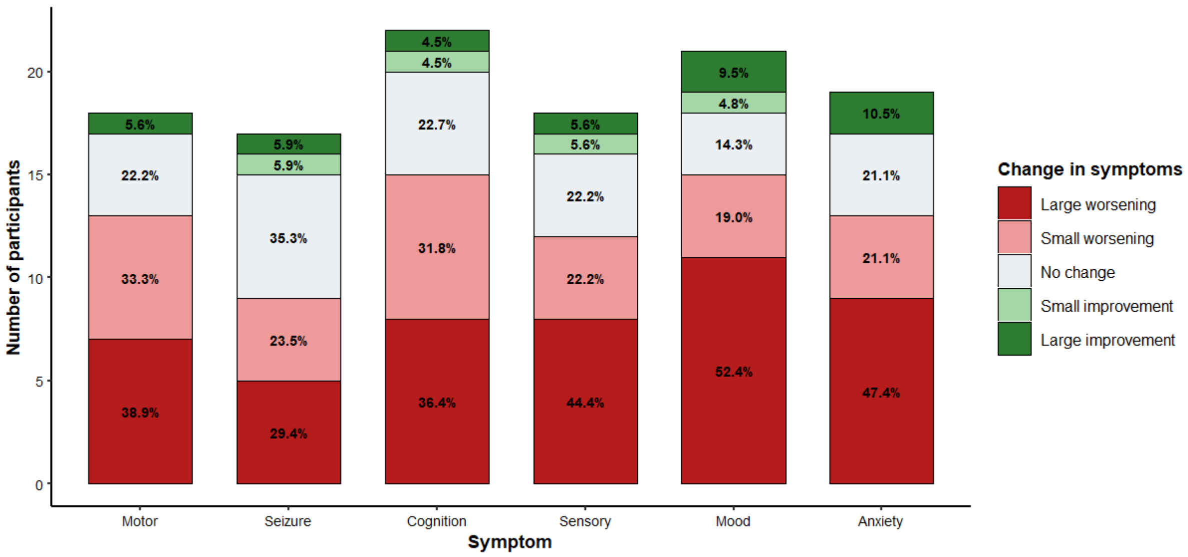


**Figure S3. Distribution of symptoms changes during menopause**

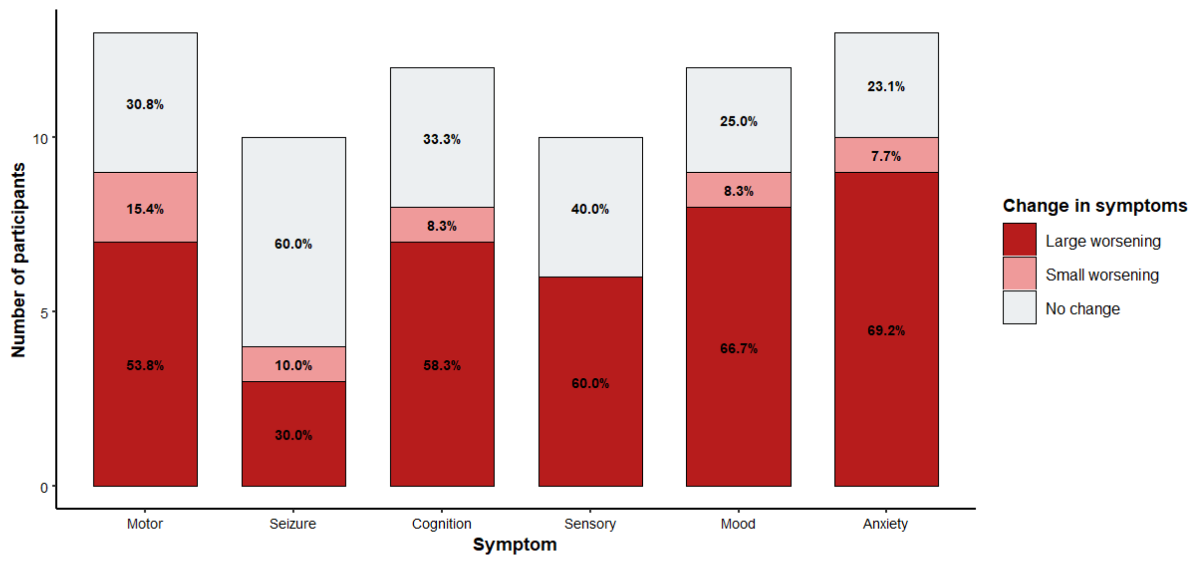


**Figure S4. Distribution of symptoms changes during perimenopause**

#### Role of hormonal replacement therapy

**Total *n* = 6**

| | |
|---|---|
| **Gender identity** | |
| Transgender female | 3 (50%) |
| Transgender male | 2 (33.3%) |
| Non-binary | 1 (16.6%) |
| **Hormones used** | |
| Estrogenes | 2 (33.3%) |
| Estrogenes and Progesterone | 1 (16.6%) |
| Testosterone | 3 (50%) |
| **Age, mean (SD)** | 24.5 (5.5) |
| **Age, minimum** | 20 |
| **Age, maximum** | 33 |
| **With motor symptoms** | 4 (66.6%) |
| **With Functional/Dissociative Seizures** | 4 (66.6%) |
| **With cognitive symptoms** | 4 (66.6%) |
| **With sensory symptoms** | 4 (66.6%) |
| **With dizziness/nausea** | 5 (83.3%) |

**Table S11. Demographic of FND participants that have been through hormonal replacement therapy (n = 6).** For participants reporting a gender transition, this table shows the distribution of age and gender, and the number of participants reporting each FND symptom within this subgroup. A participant may report multiple symptoms.

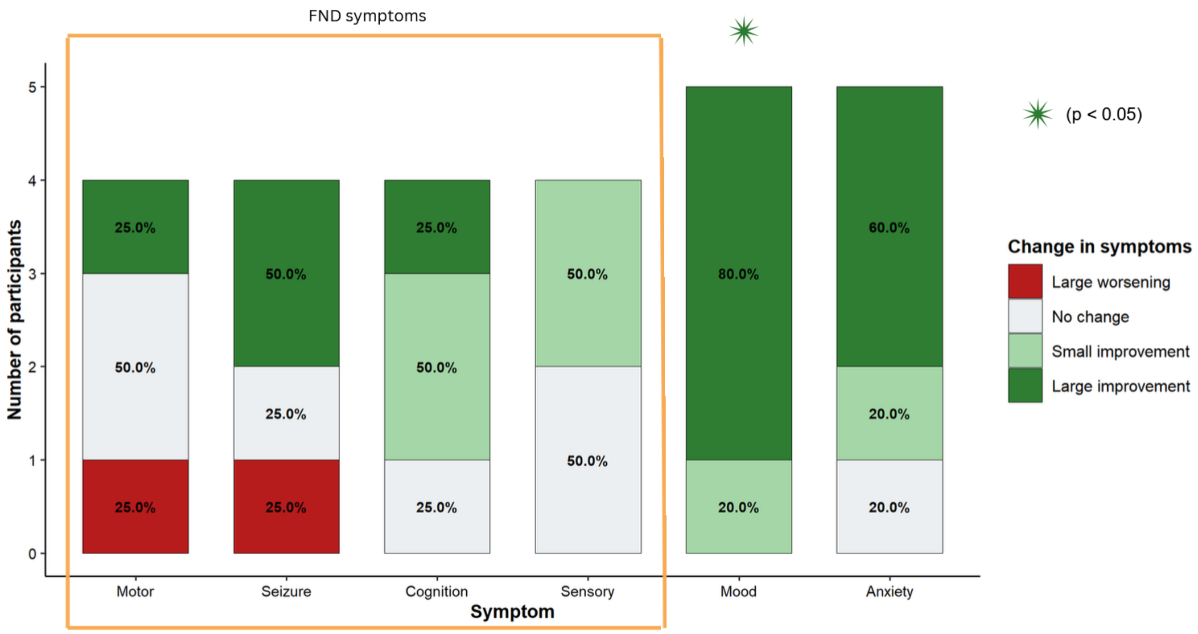


**Figure S5. Distribution of symptoms changes during gender transition by type of symptom.** The green star indicates a significant improvement in symptoms.

| Symptom | % Any Change | Large Worsening | Small Worsening | Large Improvement | Small Improvement | Mean Change |
|---|---|---|---|---|---|---|
| Motor | 50% (2) | 25% (1) | 0% (0) | 25% (1) | 0% (0) | 3 |
| Seizure | 75% (3) | 25% (1) | 0% (0) | 50% (2) | 0% (0) | 2.5 |
| Cognitive | 75% (3) | 0% (0) | 0% (0) | 25% (1) | 50% (2) | 2 |
| Sensory | 50% (2) | 0% (0) | 0% (0) | 0% (0) | 50% (2) | 2.5 |
| Vertigo | 60% (3) | 0% (0) | 0% (0) | 40% (2) | 20% (1) | 2 |
| Mood | 100% (5) | 0% (0) | 0% (0) | 80% (4) | 20% (1) | 1.2 |
| Anxiety | 80% (4) | 0% (0) | 0% (0) | 60% (3) | 20% (1) | 1.6 |

*$p < 0.05$ = significant change

**Table S12. Change in FND symptoms during gender transition (n = 6)** Among participants reporting being menopausal, % reporting any change, and direction of change (large/small improvement/worsening). Mean change score (1=large improvement → 5=large worsening) and Wilcoxon test result.

#### Regularity of the cycle across participants

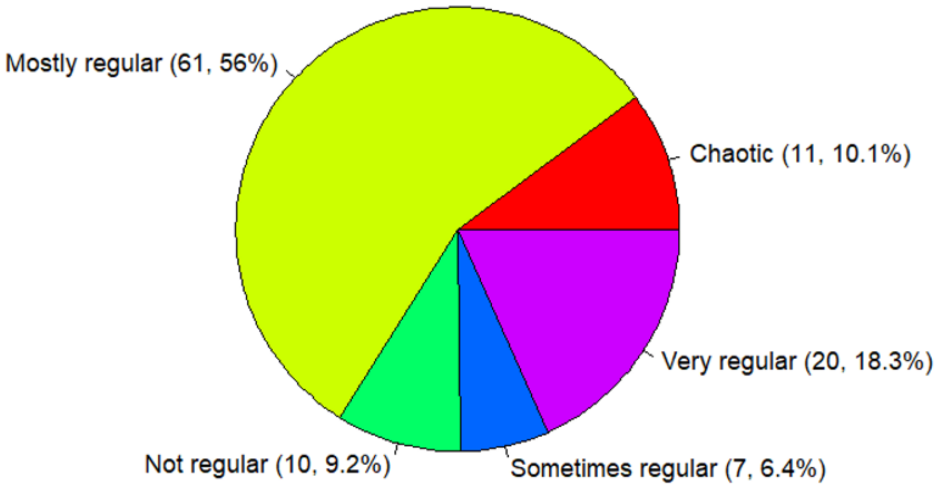


Figure S6. Distribution of cycle regularity among participants who reported a menstrual cycle

Among participants who answered having a menstrual cycle (n=88), 19.1% had a cycle "Very regular", 55.5% a cycle "Mostly regular[1] ", 6.4% "Sometimes regular", 9.1% "Not regular". For the 21 participants that indicated a cycle that was either "Sometimes regular" or "Not regular", we additionally asked whether they were diagnosed with Polycystic ovaries syndrome (PCOS). 16 of them (76.2%) reported this diagnosis.
